# Prediction of impulse control disorders in Parkinson’s disease: a longitudinal machine learning study

**DOI:** 10.1101/2025.06.04.25328442

**Authors:** Alexandros Vamvakas, Tim Van Balkom, Guido Van Wingen, Jan Booij, Daniel Weintraub, Henk W. Berendse, Odile A. van den Heuvel, Chris Vriend

## Abstract

**Background:** Impulse control disorders (ICD) in Parkinson’s disease (PD) patients mainly occur as adverse effects of dopamine replacement therapy. Despite several known risk factors associated with ICD development, this cannot yet be accurately predicted at PD diagnosis.

**Objectives:** We aimed to investigate the predictability of incident ICD by baseline measures of demographic, clinical, dopamine transporter single photon emission computed tomography (DAT-SPECT), and genetic variables.

**Methods:** We used demographic and clinical data of *medication-free* PD patients from two longitudinal datasets; Parkinson’s Progression Markers Initiative (PPMI) (n=311) and Amsterdam UMC (n=72). We extracted radiomic and latent features from DAT-SPECT. We used single nucleotic polymorphisms (SNPs) from PPMI’s NeuroX and Exome sequencing data. Four machine learning classifiers were trained on combinations of the input feature sets, to predict incident ICD at any follow-up assessment. Classification performance was measured with 10×5-fold cross-validation.

**Results:** ICD prevalence at any follow-up was 0.32. The highest performance in predicting incident ICD (AUC=0.66) was achieved by the models trained on clinical features only. Anxiety severity and age of PD onset were identified as the most important features. Performance did not improve with adding features from DAT-SPECT or SNPs. We observed significantly higher performance (AUC=0.74) when classifying patients who developed ICD within four years from diagnosis compared with those tested negative for seven or more years.

**Conclusions:** Prediction accuracy for later ICD development, at the time of PD diagnosis, is limited; however, it increases for shorter time-to-event predictions. Neither DAT-SPECT nor genetic data improve the predictability obtained using demographic and clinical variables alone.

## INTRODUCTION

Impulse control disorders (ICD) affect about 15-30% of the Parkinson’s disease (PD) population (1). Examples of ICD include hypersexuality, pathological gambling, binge eating and compulsive shopping, and are regarded as adverse effect of dopamine replacement therapy (DRT)(1). Besides DRT, various demographic (e.g., male sex, younger age of PD onset, education level), clinical (e.g., depression, sleep disorders, personal and/or family history of behavioral addictions), and genetic risk factors (2) have been associated with ICD development. So far, none of these factors are used clinically because of insufficient predictive ability beyond group-level comparisons.

Recent studies have used machine learning (ML) to predict ICD incidence at individual level (3-6). The best performing models achieved 75-85% accuracy in predicting a positive ICD screen between annual patient visits. Reported predictors across studies varied, but included demographics, disease duration, past presence of ICD, depression, antidepressant use and DRT. Additionally, two studies reported significantly higher performance when adding genetic data (3, 7), while two other studies did not (5, 6). These studies present promising evidence regarding the potential of ICD risk prediction throughout the disease trajectory. However, from a clinical perspective it is essential to predict ICD development before the initiation of DRT. This would allow personalized assessment of the premorbid vulnerability for ICD, and hence would enable clinicians to weigh the risk in deciding on DRT prescription to reduce the risk of ICD development.

ICD is associated with dysregulation of dopamine levels in the striatum. Previous imaging studies using dopamine transporter single photon emission computed tomography (DAT-SPECT), have consistently shown lower striatal DAT binding in individuals with PD and co-morbid ICD than those without ICD (8-10). However, it still remains unclear whether the observed differences are due to pre-existing neural characteristics, effects of prolonged DRT or alterations associated with ICD development (1). Nevertheless, significantly lower striatal DAT availability, measured by specific binding ratios (SBRs), was observed to precede ICD development in a small clinical sample (11). Adequately powered longitudinal studies in drug-naïve patients are needed to understand whether these changes may represent biomarkers of premorbid vulnerability to ICD (9).

Here, we aimed to develop ML models to predict future ICD development using clinical, DAT-SPECT, and genetic variables obtained from PD patients *before* the start of dopaminergic medication. We hypothesized that DAT-SPECT has added value to clinical variables in predicting ICD development. This information could be useful during the clinical management of PD.

## METHODS

### Pre-registration and reporting

This study was pre-registered at the Open Science Framework (https://osf.io/g82j3). The reporting of the prediction models adheres to the TRIPOD+AI guidelines (12) (supplemental material S1).

### Participants

#### PPMI dataset

Parkinson’s Progression Markers Initiative (PPMI) is an ongoing, longitudinal cohort study that follows individuals with PD, individuals at risk of developing PD and healthy controls (HCs) for over ten years using clinical, neuroimaging, and biological data (13). We selected a subsample that adhered to the following criteria: a) diagnosed with PD (including the pathogenic variants GBA and LRRK2, b) enrollment prior to 2015 (to increase likelihood of adequate follow-up duration), c) an abnormal baseline DAT-SPECT scan, and d) absence of a positive screen for ICD or impulse control behavior (ICB) at baseline. All participants provided written informed consent and did not use DRT at baseline.

#### Amsterdam University Medical Center (AmsterdamUMC) dataset

We enrolled patients referred to the AmsterdamUMC outpatient clinic for movement disorders that were followed-up as part of four different longitudinal studies (supplemental material S2). Participants at baseline a) were *de novo* for DRT, b) underwent a DAT-SPECT scan, and c) were free of ICD/ICB. All participants provided written informed consent, obtained according to the Declaration of Helsinki, and the study protocol was reviewed and approved by the Medical Ethical Committee.

### Impulse control disorder classification

ICD/ICB was screened using the questionnaire for ICD (QUIP) and the QUIP-rating scale (QUIP-RS), for PPMI and AmsterdamUMC samples, respectively, using the published optimal cut-offs (14-16). Individuals with PD were classified as ICD positive if they scored above the cut-off for hypersexuality, binge eating, pathological gambling, or compulsive buying at any follow-up time-point. Individuals that screened positive for ICB (punding, hobbyism, and walkabouts (for the QUIP) or dopamine dysregulation syndrome (for the QUIP-RS)) but not for ICD, were excluded from the analysis. Individuals were classified as no-ICD if they screened negative across all time-points.

### Clinical measurements

Clinical measurements and employed harmonization steps across the PPMI and AmsterdamUMC samples are outlined in supplemental Table S3. We used information for disease stage, motor symptom severity, severity of depression, anxiety and apathy symptoms, and global cognitive function. An REM sleep behavioral disorder (RBD) screener was available only for the PPMI sample. We calculated the levodopa-equivalent daily dose (LEDD) at each time-point as earlier described (17).

### DAT-SPECT image acquisition

In both samples presynaptic striatal DAT binding was measure with [^123^I]FP-CIT-SPECT, according to the European Association of Nuclear Medicine procedure guidelines (18). We obtained the filtered reconstructed [^123^I]FP-CIT-SPECT volumes from their original sources. Details regarding the imaging protocols, reconstruction and processing are given in supplemental material S4.

### Image post-acquisition processing and segmentation

We spatially normalized the DAT-SPECT volumes to Montreal Neurologic Institute coordinates with SPM12 (https://www.fil.ion.ucl.ac.uk/spm/software/spm12/) using a previously published template (19). A histogram matching method (20), was utilized to normalize voxel intensities to reduce inter-subject/site differences. We delineated eight volumes of interest (VOIs), namely ventral striatum, anterior putamen, posterior putamen and caudate nucleus (all left and right) using FSL’s Structural Striatal Atlas (21). As pre-registered, thalamic ROIs were also segmented with FSL atlas, but they were not used in the analysis due to very low signal-to-noise ratio. The cerebellum was manually delineated and used as reference region (representing non-specific binding) in SBRs calculation.

### Specific binding ratios, radiomics and deep learning (latent) features extraction

We defined the striatal SBR using Equation 1;

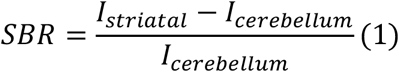

where, *I* is the average voxel intensity. We used the Pyradiomics library (22) to extract 1232 radiomic features from each striatal VOI, considering the original, as well as derived volumes using Wavelet decompositions and laplacian of gaussian filters. Additionally, we implemented a 3D autoencoder in Tensorflow/Keras 2.0 library (keras.io), adjusting the architecture previously described in (23).This was used to extract latent features from cropped striatum VOIs of size 32×32×24 voxels. Feature extraction details are given in supplemental material S5.

### Genetic information

PPMI genetic data were originally obtained with NeuroX genotyping array and whole Exome sequencing. We used the PLINK 1.9 software (https://www.cog-genomics.org/plink/) to extract 16 single nucleotic polymorphisms (SNPs) of 13 genes, previously associated with increased risk of ICD development (2-4, 6) (supplemental Table S10).

### Machine Learning

We trained logistic regression (LR), random forest (RF), gradient boosting (GB) and multilayer perceptron (MLP) classifiers with scikit-learn library (https://scikit-learn.org/stable/) in Python (version 3.9), using the clinical features subset alone, the clinical+radiomic features subset and the clinical+latent features subset. Two additional models considering clinical+genetic, and clinical+genetic+radiomic features were evaluated on the PPMI dataset only, as the AmsterdamUMC sample did not include genetic data. We excluded participants with missing SNP data. Missing values of clinical features were imputed with scikit-learn iterative imputer. All input values were standardized to zero mean and unit variance. We employed two-step feature selection to keep a smaller subset of radiomic features. First, we reduced the dimensionality of the radiomic feature space to a predefined number of 50 features using the minimum redundance-maximum relevance algorithm. We subsequently identified the optimal feature subset using the least absolute shrinkage and selection operator (LASSO). Models were tested with stratified 5-fold cross-validation, repeated 10 times. Feature imputation, standardization and selection transformations were separately learnt from training and applied to testing dataset splits. Nested 5-fold cross-validations and grid-search were utilized for hyperparameter tuning. Classification performance was assessed with balanced accuracy, sensitivity, specificity, and receiver operating characteristic area under curve (AUC). Permutation feature importances (30 permutations), were calculated to measure the contribution of each input feature to the fitted models’ statistical performance, with the mean decrease in accuracy (MDA) metric. An overview of the methodology is depicted in Figure 1.

**Figure 1.**
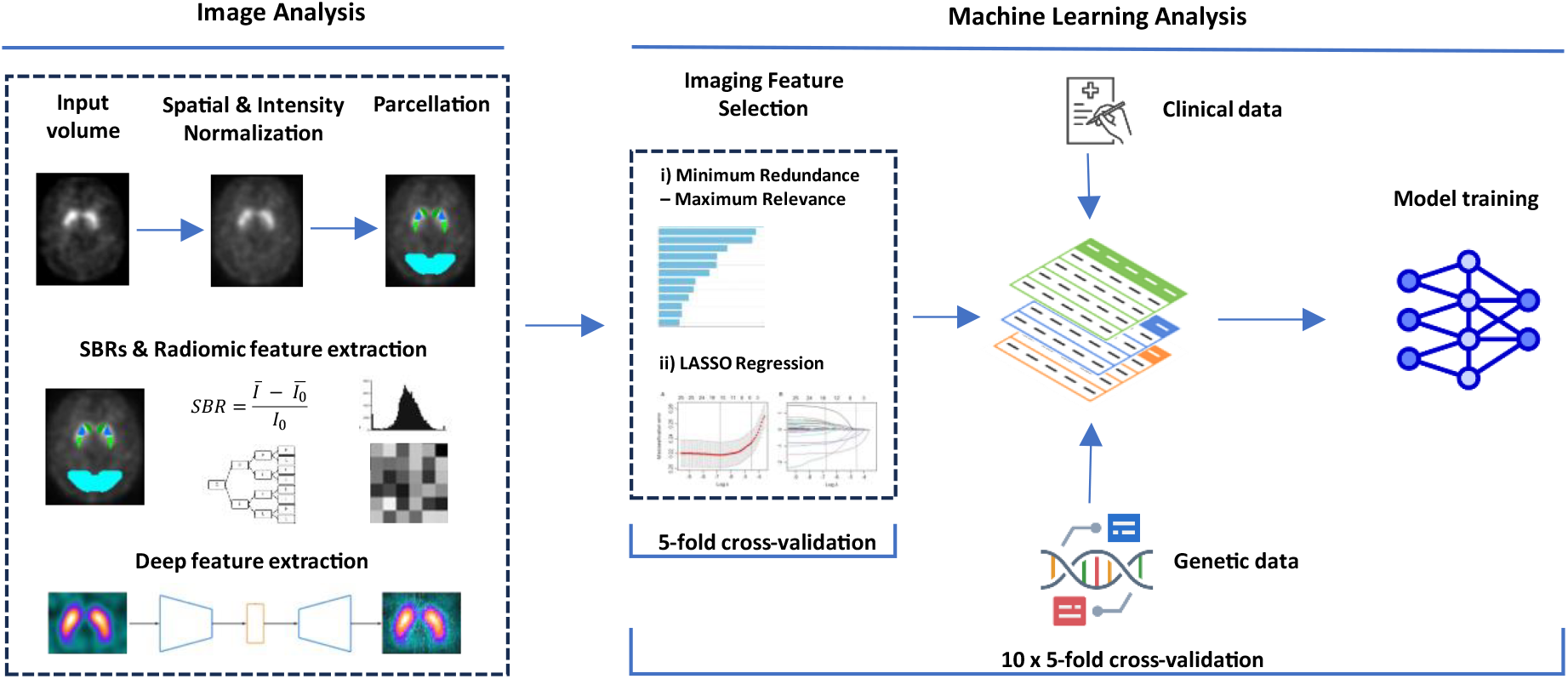
The overall study workflow. DAT-SPECTs were spatially and intensity normalized. 3D striatum parcellation to different regions and a whole-striatum patch were used for radiomic and latent extraction, respectively. Selected radiomic features by the nested pipeline were fused with clinical and genetic data for model development with a repeated 5-fold cross-validation scheme.

### Statistical Analyses

Statistical analyses were performed in R (version 4.3.2; 24). We performed univariate independent samples t-tests, to compare demographic and clinical information of the PPMI and AmsterdamUMC datasets. For skewed distributions and categorical variables, we used Mann-Whitney or Fisher’s exact tests, respectively. To assess differences between ICD vs No-ICD groups, we used linear/logistic regression with the clinical features or SBRs as dependent variable and group as fixed factor. We additionally performed linear regression including cohort and its interaction with group as covariate of interest, and age, sex and education level as additional covariates. We corrected for multiple comparisons using the Benjamini-Hochberg false discovery rate (FDR) (25). We tested ML models’ statistical significance against chance with permutation testing. We performed pairwise comparisons of classification performances across models and feature subsets either using the paired-samples t-tests, in case of identical train-test splits, corrected for repeated k-fold cross-validation (26, 27), or the Welch’s t-test in case of non-identical train-test splits. All significance levels were set to a=0.05.

## RESULTS

### Participants

The final sample consisted of 378 patients (PPMI n=311, AmsterdamUMC n=67; Figure 2). Disease severity at baseline was higher in the AmsterdamUMC sample for both motor and non-motor symptoms, and the follow-up duration was higher in the PPMI sample. The prevalence of ICD at any follow-up was 32.0% (PPMI: 32.5%, AmsterdamUMC: 29.9%) (supplemental Table S6).

**Figure 2.**
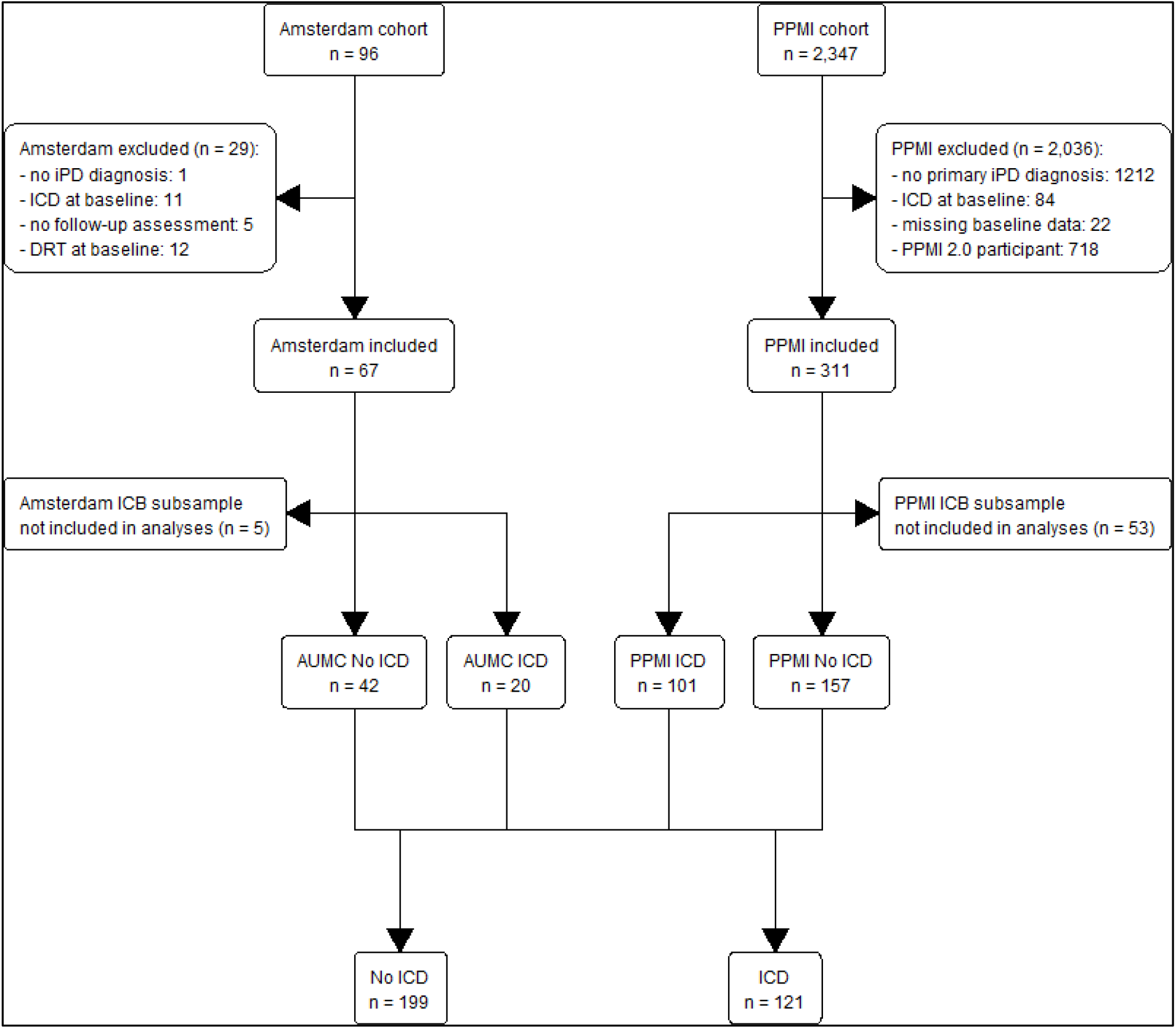
Flowchart with reasons for exclusion and ICD group classification per cohort and in total. *Abbreviations:* ICB = impulsive compulsive behaviors; ICD = impulse control disorder; DRT = dopamine replacement therapy; iPD = idiopathic Parkinson’s disease.

### Univariate group analyses

The ICD group had a lower age at onset and a higher proportion of severe apathy symptoms at baseline, compared with the No ICD group. The ICD group showed a lower LEDD score at follow-up, with a lower levodopa dosage but a significantly higher dopamine agonist dosage. The lower age at onset and lower LEDD at follow-up were specific to the PPMI sample when inspecting the Cohort*Group interaction. Moreover, in the AmsterdamUMC cohort the ICD group showed higher MDS-UPDRS part III scores, lower MMSE score, and higher depressive, anxiety and apathy symptom severity. Group statistics are shown in Table 1 and supplemental Table S7. Univariate test statistics are shown in supplemental Table S8. There were no significant differences between the No ICD and ICD group on specific binding ratio’s, radiomic texture features and genetic features after controlling for FDR (supplemental Tables S9-S11).

**Table 1.**
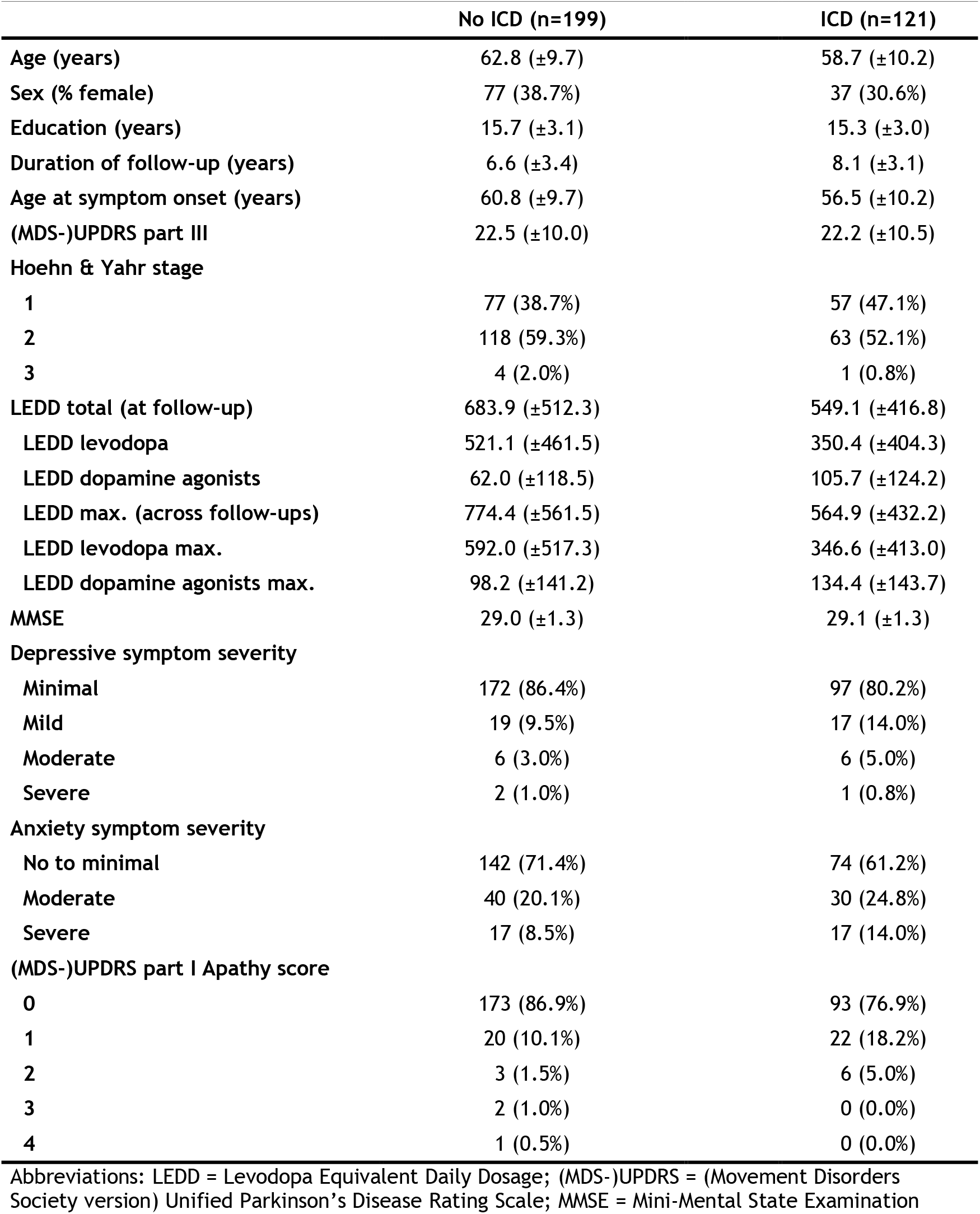
Group statistics of the ICD, No ICD groups in the full combined dataset (PPMI+AmsterdamUMC).

### Pre-registered ML analyses

Cross-validated classification metrics of the different models on the combined dataset (PPMI+ AmsterdamUMC) are presented in Table 2. For the models including only clinical variables, the RF, LR and MLP models were significant against chance (AUC=0.66±0.05, p<0.05), while GB did not (AUC=0.57±0.06, p=0.86). The most important predictor of ICD were (higher) anxiety and (lower) age of onset, with MDA=0.07±0.05 and MDA=0.04±0.04, respectively (Figure 3). Model performance dropped considerably when adding DAT-SPECT features (RF model with radiomic features: AUC=0.60±0.06, LR and MLP classifiers with latent features: AUC=0.63±0.06). Anxiety, age of onset and three radiomic features showed equally high permutation importance (MAD=0.03±0.04) (Figure 3).

**Table 2.** Cross-validated performance evaluation of ICD vs non-ICD classification models in the full combined dataset (PPMI+ AmsterdamUMC)

| Feature subset | Model | Accuracy | Balanced accuracy | Sensitivity | Specificity | AUC |
| --- | --- | --- | --- | --- | --- | --- |
| Clinical features | RF | $0.62 \pm 0.05$ | $0.62 \pm 0.05$ | $0.61 \pm 0.10$ | $0.62 \pm 0.08$ | $0.66 \pm 0.05$ |
| | LR | $0.63 \pm 0.06$ | $0.62 \pm 0.06$ | $0.59 \pm 0.09$ | $0.65 \pm 0.07$ | $0.66 \pm 0.06$ |
| | GB | $0.58 \pm 0.06$ | $0.55 \pm 0.06$ | $0.42 \pm 0.15$ | $0.67 \pm 0.08$ | $0.57 \pm 0.06$ |
| | MLP | $0.65 \pm 0.05$ | $0.58 \pm 0.05$ | $0.30 \pm 0.09$ | $0.86 \pm 0.08$ | $0.66 \pm 0.05$ |
| Clinical + Radiomic features | RF | $0.60 \pm 0.06$ | $0.58 \pm 0.06$ | $0.52 \pm 0.10$ | $0.64 \pm 0.08$ | $0.60 \pm 0.06$ |
| | LR | $0.57 \pm 0.05$ | $0.56 \pm 0.05$ | $0.52 \pm 0.09$ | $0.60 \pm 0.08$ | $0.59 \pm 0.05$ |
| | GB | $0.57 \pm 0.07$ | $0.54 \pm 0.06$ | $0.41 \pm 0.11$ | $0.67 \pm 0.09$ | $0.56 \pm 0.07$ |
| | MLP | $0.58 \pm 0.07$ | $0.52 \pm 0.05$ | $0.28 \pm 0.19$ | $0.77 \pm 0.20$ | $0.55 \pm 0.06$ |
| Clinical + Latent features* | RF | $0.61 \pm 0.06$ | $0.59 \pm 0.06$ | $0.49 \pm 0.10$ | $0.68 \pm 0.10$ | $0.61 \pm 0.06$ |
| | LR | $0.60 \pm 0.06$ | $0.59 \pm 0.06$ | $0.56 \pm 0.09$ | $0.63 \pm 0.08$ | $0.63 \pm 0.06$ |
| | GB | $0.57 \pm 0.06$ | $0.54 \pm 0.06$ | $0.40 \pm 0.11$ | $0.68 \pm 0.08$ | $0.56 \pm 0.06$ |
| | MLP | $0.63 \pm 0.06$ | $0.56 \pm 0.06$ | $0.31 \pm 0.14$ | $0.82 \pm 0.12$ | $0.63 \pm 0.06$ |

Abbreviations: AUC = Area Under Curve, RF = Random Forest, LR = Logistic Regression, GB = Gradient Boosting, MLP = Multi-Layer Perceptron. \*Latent features were extracted with the 3D autoencoder network.

**Figure 3.**
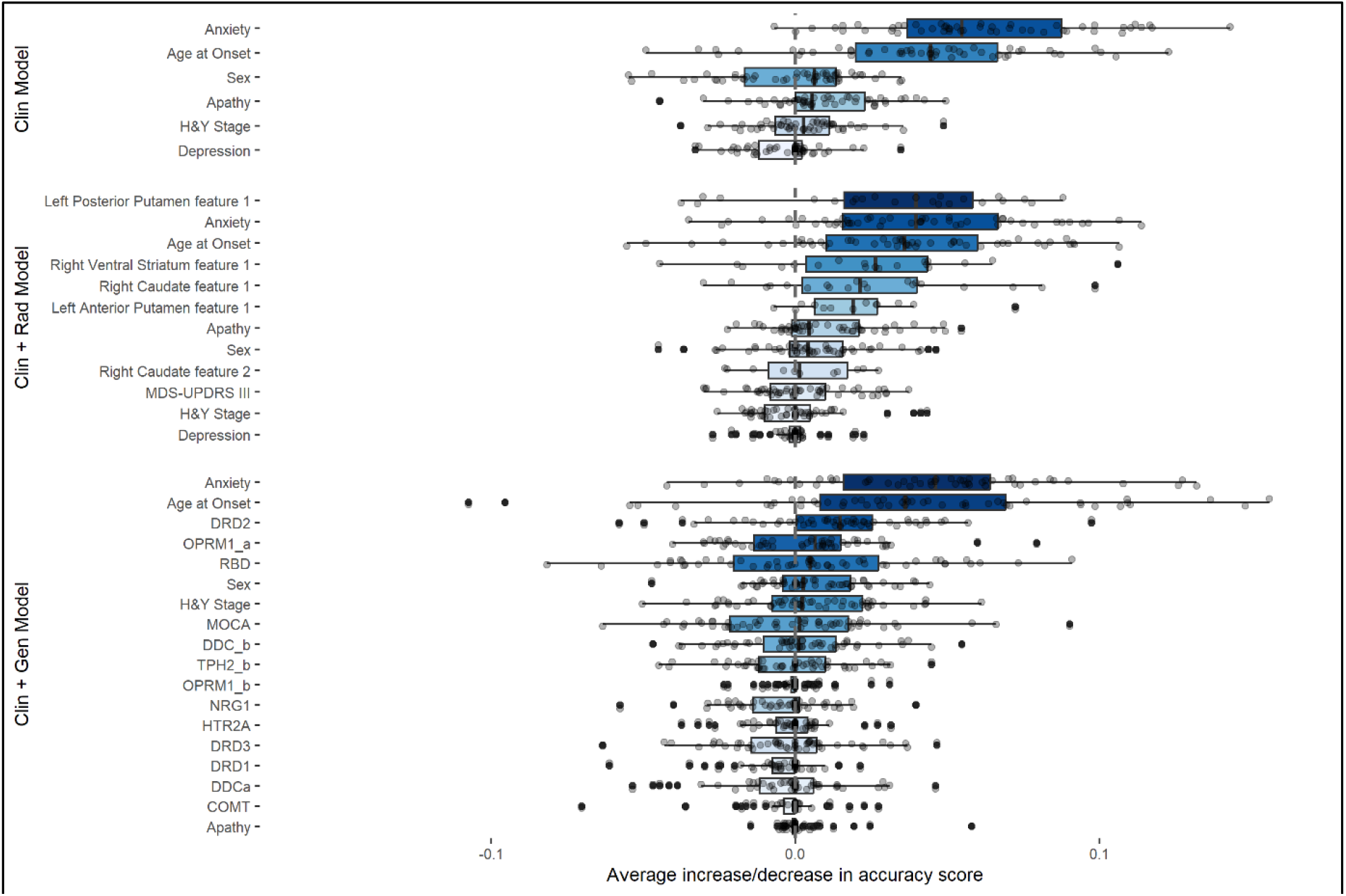
Permutation feature importances for the clinical (Top), clinical + radiomics (Middle) and the clinical + genetics (Bottom; PPMI only) RF models.

Results of the models with additional inclusion of SNPs for the PPMI cohort are shown in Table 3. Highest performance was achieved by the RF classifier, however no significant improvement was observed with the addition of genetic data (AUC=0.69±0.06) to the Clinical (AUC=0.68±0.06) or Clinical+Radiomics (AUC=0.69±0.06) models. The highest feature importance was observed for anxiety (MDA=0.08±0.05) and age of onset (MDA=0.06±0.05). Two radiomic features presented positive MDA=0.01±0.04, while for the other selected features and the genetic variants MDA was zero or negative. (Figure 3).

**Table 3.** Cross-validated performance evaluation of ICD vs non-ICD classification models in the PPMI dataset.

| Feature subset | Model | Accuracy | Balanced accuracy | Sensitivity | Specificity | AUC |
| --- | --- | --- | --- | --- | --- | --- |
| Clinical features | RF | 0.64 ± 0.06 | 0.64 ± 0.06 | 0.65 ± 0.12 | 0.63 ± 0.10 | 0.68 ± 0.06 |
|  | LR | 0.63 ± 0.07 | 0.63 ± 0.07 | 0.62 ± 0.14 | 0.64 ± 0.08 | 0.67 ± 0.07 |
|  | GB | 0.57 ± 0.07 | 0.55 ± 0.07 | 0.44 ± 0.11 | 0.66 ± 0.10 | 0.58 ± 0.07 |
|  | MLP | 0.57 ± 0.08 | 0.56 ± 0.08 | 0.41 ± 0.12 | 0.71 ± 0.15 | 0.60 ± 0.08 |
| Clinical + Genetics features | RF | 0.65 ± 0.06 | 0.64 ± 0.06 | 0.61 ± 0.12 | 0.67 ± 0.09 | 0.69 ± 0.06 |
|  | LR | 0.60 ± 0.07 | 0.60 ± 0.07 | 0.58 ± 0.11 | 0.61 ± 0.08 | 0.64 ± 0.07 |
|  | GB | 0.61 ± 0.06 | 0.58 ± 0.07 | 0.47 ± 0.13 | 0.69 ± 0.09 | 0.62 ± 0.07 |
|  | MLP | 0.63 ± 0.06 | 0.59 ± 0.07 | 0.42 ± 0.13 | 0.75 ± 0.09 | 0.63 ± 0.07 |
| Clinical + Genetics + Radiomics features | RF | 0.65 ± 0.05 | 0.63 ± 0.06 | 0.55 ± 0.12 | 0.72 ± 0.08 | 0.69 ± 0.06 |
|  | LR | 0.63 ± 0.06 | 0.62 ± 0.06 | 0.58 ± 0.11 | 0.66 ± 0.10 | 0.67 ± 0.06 |
|  | GB | 0.65 ± 0.05 | 0.61 ± 0.06 | 0.48 ± 0.13 | 0.74 ± 0.08 | 0.68 ± 0.06 |
|  | MLP | 0.59 ± 0.09 | 0.56 ± 0.08 | 0.42 ± 0.25 | 0.70 ± 0.22 | 0.61 ± 0.08 |

Abbreviations: AUC = Area Under Curve, RF = Random Forest, LR = Logistic Regression, GB = Gradient Boosting, MLP = Multi-Layer Perceptron

Performance of predicting positive screens for specific ICDs vs No-ICD with models trained on clinical features were: gambling; AUC=0.57±0.17 (p=0.985), hypersexuality; AUC=0.74±0.06 (p=0.001), compulsive buying; AUC=0.62±0.09 (p=0.005), binge eating; AUC=0.67±0.09 (p=0.004). No improvement was observed with the addition of imaging features. Notably, sex was the most important predictor of hypersexuality (MDA=0.12±0.04). Anxiety (MDA=0.8±0.07) followed by age of onset (MDA=0.6±0.05) were important predictors for the other ICDs, while PD stage presented some importance in binge eating prediction (MDA=0.3±0.05).

### Post-hoc ML analyses

#### Predicting ICD up to the 4th-year follow-up

As the prediction of an event further into the future may be more difficult, we performed a post-hoc analysis including ICD positive patients that had developed ICD *within 4 years* after baseline, while those that developed ICD after 4 years were removed from analyses. We contrasted this subgroup with patients that did not show any positive screen for 7 or more years after baseline. All models achieved significantly higher performances (p<0.05) compared to the pre-registered analysis, up to AUC=0.74±0.05.

#### Adding medication use at time of ICD development as predictor

Since DRT is considered the primary risk factor of ICD development we additionally included medication received at the time of ICD positive screening as predictor (LEDD at follow-up, as well as usage of levodopa, dopamine agonists, MAO-B and COMT inhibitors (yes or no)). All models achieved significantly higher performances compared to the models of the pre-registered analysis (p<0.05), up to AUC=0.71±0.06, with LEDD at follow-up being important (MDA=0.02±0.04), along with age at onset (MDA=0.04±0.04) and anxiety (MDA=0.03±0.04).

#### Machine learning diagnosis of PD vs Healthy Controls

Lastly, we evaluated ML models to classify PD vs Healthy Controls to confirm the quality of the extracted radiomic and latent features. We used the imaging data of the 378 PD samples of the pre-registered analysis and the 180 HCs of the PPMI. The models achieved excellent performance of AUC=0.99±0.02 (supplemental material S12).

## DISCUSSION

In this study we sought to investigate the ability of ML models, trained on clinical, DAT-SPECT and genetic data obtained at the time of PD diagnosis, to predict the future development of ICDs after starting DRT. We observed a moderate performance of the preregistered models using baseline clinical variables in predicting ICD positive screen at any time follow-up assessment (AUC=0.66±0.05). The most important predictors were the severity of anxiety and age of PD onset. Contrary to our hypothesis, there was no improvement in the predictive performance of the models when either DAT-SPECT features or SNPs were added. In an effort to increase the relevance of our results we performed two post-hoc analyses. First, we used a modified sample including patients who developed ICD within 4 years from diagnosis and those who tested negative for seven or more years, which boosted the performance up to AUC=0.74±0.05. Additionally we tested adding dopaminergic medication use at the time of ICD development as predictor variable, which also significantly increased the performance up to AUC=0.71±0.06.

The most important contribution of our study to the existing knowledge is the evaluation of molecular neuroimaging in the prediction of future ICD development, at the time of PD diagnosis and before DRT initiation. Previous DAT-SPECT studies have identified significantly lower striatal DAT binding in PD patients with ICD at the group level (8, 9, 11). However, whether this reduction is a consequence of either DRT or ICD, or predates ICD development, and thus could be used as a predictive biomarker on the individual level remained unclear. Here we extracted radiomic features, that were previously shown to be more sensitively associated with future PD outcomes, compared to the conventional SBRs (28, 29), but these did not improve the performance of the ML model, showing negligible predictive ability of DAT-SPECT at the individual level. The same trend was also observed with the models including latent features derived from the autoencoder. We did however observe excellent classification accuracy (AUC=0.99±0.02) when trying to classify PD versus healthy controls, meaning that the extracted features contain relevant information about the PD pathophysiology.

Recent studies have highlighted the promise of ML to predict ICD incidence at the individual level (3-6), albeit showcasing a great variability in the methodology used (PD sample, predictor and target variables, validation setting) and, consequently, with mode performances ranging between AUCs=0.65-0.85. In our study we observed a classification performance AUC=0.66 for the clinical features subset, that lies within the lower boundary of the previously reported performances. However, there are major methodological differences between this and the previous studies. First, previous studies used data from all available assessments up to the point of ICD development for their models. In contrast, we aimed at assessing the predictive performance of data obtained around the time of PD diagnosis, and this might have reduced the predictive effect, especially when the time interval to ICD development is greater. Only Faouzi et al. (5) have reported a baseline model with AUC=0.75. However, this model had presence of ICD at baseline as the most important predictor, while we excluded PD cases with a positive ICD screen at baseline from the analyses, as these are likely false positives as no DRT had commenced. Second, previous studies have also used DRT as predictor, which turned out to be the most significant predictor, in line with a wealth of previous evidence that have directly linked ICD incidence to DRT (1, 30). Indeed, we were able to replicate these findings when adding DRT use at the time of ICD development as predictor. However, we do not consider these models useful in terms of their clinical applicability, since they would rely on the knowledge of future data beforehand.

In our study, we did not observe any contribution from the genetic data to the models. Previous studies have provided mixed results regarding the predictive value of SNPs for ICD development. Erga et al. (7) and Jesus et al. (31) reported significant improvements when adding genetic features to their ML models, trained on the Norwegian ParkWest study sample and a local Spanish dataset, respectively. In contrast, Redenŝek et al. (6) did not when using a local Slovenian PD sample. Kraemmer and colleagues (3) reported an AUC of 0.65 with clinical variables only, and an AUC of 0.76 with clinical features and 13 SNPs, denoting a significant improvement in ICD prediction, in the PPMI dataset. However, none of these SNPs where found significant in their follow-up study (4), which also included samples from the 23&Me, and a local UPenn dataset. Although the latter study (4) achieved relatively high classification performance (AUC=0.72), the two SNPs included in their model were not significant predictors of ICD. Faouzi et al. (5) who included 50 SNPs previously proposed as risk factors of ICD, did not observe a significant improvement in the predictive performance of their models.

This is the first study that combined multiparametric data obtained around the time of PD diagnosis to predict ICD development before initiation of DRT. In the majority of the ML models, the severity of anxiety symptoms was the most important predictor. This finding supports previous evidence showing higher anxiety levels preceding ICD development (31, 32). Neuropsychiatric symptom severity is also reported to be associated with future cognitive and motor decline (33, 34). These findings underscore the importance of neuropsychiatric screening at PD diagnosis, to adequately diagnose and manage ICD symptom development. However, our study shows that presence of anxiety—whether or not combined with other clinical, DAT-SPECT or genetic information—is not sufficient to predict ICD development at the individual level. As observed in this and previous studies (5), higher accuracy could be achieved when predicting closer to ICD onset, and this also might be of clinical importance.

Recent reports demonstrated significant differences between ICD positive and negative PD patients in magnetic resonance imaging (MRI) measures, such as decreased functional connectivity within and between dopaminergic neuronal circuitries, periventricular white matter hyperintensities and widespread white matter tract damage, and aberrant brain topological organization (35-37). Such measures might be of interest to include in future prediction studies.

ML modelling needs adequate sample sizes to achieve good predictive performance, thus our results might have been influenced by the relatively low sample size. Although the inclusion of different datasets is vital, this has increased the heterogeneity in two ways here. First the heterogeneity in the clinical status of PD, which is a chronic disease with variable progression pathways, as the AmsterdamUMC sample includes more progressed cases compared to PPMI, and second, with the variability in the assessments/examinations between sites. An additional limitation is that fewer follow-up assessments were obtained for the AmsterdamUMC sample.

In conclusion, our study shows that clinical variables, and presence of anxiety in particular, of DRT-naive PD patients might indicate vulnerability for future ICD development, but these are not sufficient to provide accurate predictions at level of the individual patient to optimize the clinical management. Neither DAT-SPECT or genetic data improved the predictability over demographic and clinical variables.

## AUTHOR ROLES

**AV:** Data curation, Investigation, Methodology, Image processing, Image analysis, Machine Learning analysis, Writing original draft. **TvB:** Data curation, Investigation, Univariate Analysis, Writing original draft, Writing review & editing. **GvW, JB, DW, HB, OvdH:** Validation, Writing review & editing, **CV:** Conceptualization, Funding acquisition, Investigation, Methodology, Validation, Writing review & editing, Project administration, Supervision.

## DATA AVAILABILITY

Data used in the preparation of this article was obtained on 1-9-2023 from the PPMI database (www.ppmi-info.org/access-dataspecimens/download-data), RRID:SCR_006431. For up-to-date information on the study, visit www.ppmi-info.org. The AmsterdamUMC dataset is not publicly available according to GDPR. Codes are available at www.github.com/sciqd/Learn_2_control

## ABBREVIATIONS

AmsterdaUMC: Amsterdam University Medical Center
AUC: Area Under Curve
DAT: Dopamine Transporter
DRT: Dopamine Replacement Therapy
FDR: False Discovery Rate
GB: Gradient Boosting
ICD/B: Impulse Control Disorders/Behaviors
iPD: idiopathic Parkinson’s Disease
LASSO: Least Absolute Shrinkage and Selection Operator
LEDD: Levodopa Equivalent Daily Dose
LR: Logistic Regression
MDA: Mean Decrease Accuracy
ML: Machine Learning
MLP: Multi-layer Perceptron
MMSE: Mini-Mental State Examination
PPMI: Parkinson’s Progression Markers Initiative
QUIP: Questionnaire for ICD in PD
RBD: Sleep Behavioral Disorder
RF: Random Forest
SBR: Specific Binding Ratio
SNP: Single Nucleotic Polymorphism
SPECT: Single Photon Emission Computed Tomography
UPDRS: Unified Parkinson’s Disease Rating Scale
VOI: Volume of Interest

## REFERENCES

1. Vriend C. The neurobiology of impulse control disorders in Parkinson’s disease: from neurotransmitters to neural networks. Cell Tissue Res. 2018;373(1):327–36.

2. Dulski J, Uitti RJ, Ross OA, Wszolek ZK. Genetic architecture of Parkinson’s disease subtypes - Review of the literature. Front Aging Neurosci. 2022;14:1023574.

3. Kraemmer J, Smith K, Weintraub D, Guillemot V, Nalls MA, Cormier-Dequaire F, et al. Clinical- genetic model predicts incident impulse control disorders in Parkinson’s disease. J Neurol Neurosurg Psychiatry. 2016;87(10):1106–11.

4. Weintraub D, Posavi M, Fontanillas P, Tropea TF, Mamikonyan E, Suh E, et al. Genetic prediction of impulse control disorders in Parkinson’s disease. Ann Clin Transl Neurol. 2022;9(7):936–49.

5. Faouzi J, Bekadar S, Artaud F, Elbaz A, Mangone G, Colliot O, et al. Machine Learning-Based Prediction of Impulse Control Disorders in Parkinson’s Disease From Clinical and Genetic Data. IEEE Open J Eng Med Biol. 2022;3:96–107.

6. Redenšek S, Jenko Bizjan B, Trošt M, Dolžan V. Clinical and Clinical-Pharmacogenetic Models for Prediction of the Most Common Psychiatric Complications Due to Dopaminergic Treatment in Parkinson’s Disease. Int J Neuropsychopharmacol. 2020;23(8):496–504.

7. Erga AH, Dalen I, Ushakova A, Chung J, Tzoulis C, Tysnes OB, et al. Dopaminergic and Opioid Pathways Associated with Impulse Control Disorders in Parkinson’s Disease. Frontiers in neurology. 2018;9:109.

8. Navalpotro-Gomez I, Dacosta-Aguayo R, Molinet-Dronda F, Martin-Bastida A, Botas-Peñin A, Jimenez-Urbieta H, et al. Nigrostriatal dopamine transporter availability, and its metabolic and clinical correlates in Parkinson’s disease patients with impulse control disorders. Eur J Nucl Med Mol Imaging. 2019;46(10):2065–76.

9. Martini A, Dal Lago D, Edelstyn NMJ, Salgarello M, Lugoboni F, Tamburin S. Dopaminergic Neurotransmission in Patients With Parkinson’s Disease and Impulse Control Disorders: A Systematic Review and Meta-Analysis of PET and SPECT Studies. Frontiers in neurology. 2018;9:1018.

10. Smith KM, Xie SX, Weintraub D. Incident impulse control disorder symptoms and dopamine transporter imaging in Parkinson disease. J Neurol Neurosurg Psychiatry. 2016;87(8):864–70.

11. Vriend C, Nordbeck AH, Booij J, van der Werf YD, Pattij T, Voorn P, et al. Reduced dopamine transporter binding predates impulse control disorders in Parkinson’s disease. Mov Disord. 2014;29(7):904–11.

12. Collins GS, Moons KGM, Dhiman P, Riley RD, Beam AL, Van Calster B, et al. TRIPOD+AI statement: updated guidance for reporting clinical prediction models that use regression or machine learning methods. BMJ. 2024;385:e078378.

13. Initiative PPM. The Parkinson Progression Marker Initiative (PPMI). Prog Neurobiol. 2011;95(4):629–35.

14. Weintraub D, Hoops S, Shea JA, Lyons KE, Pahwa R, Driver-Dunckley ED, et al. Validation of the questionnaire for impulsive-compulsive disorders in Parkinson’s disease. Mov Disord. 2009;24(10):1461–7.

15. Weintraub D, Mamikonyan E, Papay K, Shea JA, Xie SX, Siderowf A. Questionnaire for Impulsive-Compulsive Disorders in Parkinson’s Disease-Rating Scale. Mov Disord. 2012;27(2):242–7.

16. Evans AH, Okai D, Weintraub D, Lim SY, O’Sullivan SS, Voon V, et al. Scales to assess impulsive and compulsive behaviors in Parkinson’s disease: Critique and recommendations. Mov Disord. 2019;34(6):791–8.

17. Jost ST, Kaldenbach MA, Antonini A, Martinez-Martin P, Timmermann L, Odin P, et al. Levodopa Dose Equivalency in Parkinson’s Disease: Updated Systematic Review and Proposals. Mov Disord. 2023;38(7):1236–52.

18. Darcourt J, Booij J, Tatsch K, Varrone A, Vander Borght T, Kapucu OL, et al. EANM procedure guidelines for brain neurotransmission SPECT using (123)I-labelled dopamine transporter ligands, version 2. Eur J Nucl Med Mol Imaging. 2010;37(2):443–50.

19. García-Gómez FJ, García-Solís D, Luis-Simón FJ, Marín-Oyaga VA, Carrillo F, Mir P, et al. [Elaboration of the SPM template for the standardization of SPECT images with 123I-Ioflupane]. Rev Esp Med Nucl Imagen Mol. 2013;32(6):350–6.

20. Salas-Gonzalez D, Górriz JM, Ramírez J, Illán IA, Lang EW. Linear intensity normalization of FP-CIT SPECT brain images using the α-stable distribution. Neuroimage. 2013;65:449–55.

21. Tziortzi AC, Searle GE, Tzimopoulou S, Salinas C, Beaver JD, Jenkinson M, et al. Imaging dopamine receptors in humans with [11C]-(+)-PHNO: dissection of D3 signal and anatomy. Neuroimage. 2011;54(1):264–77.

22. van Griethuysen JJM, Fedorov A, Parmar C, Hosny A, Aucoin N, Narayan V, et al. Computational Radiomics System to Decode the Radiographic Phenotype. Cancer Res. 2017;77(21):e104–e7.

23. Hosseinzadeh M, Gorji A, Fathi Jouzdani A, Rezaeijo SM, Rahmim A, Salmanpour MR. Prediction of Cognitive Decline in Parkinson’s Disease Using Clinical and DAT SPECT Imaging Features, and Hybrid Machine Learning Systems. Diagnostics (Basel). 2023;13(10).

24. R Core Team. R: A language and environment for statistical computing: R Foundation for Statistical Computing, Vienna, Austria; 2021 [Available from: https://www.R-project.org/.

25. Benjamini Y, Hochberg Y. Controlling the false discovery rate: a practical and powerful approach to multiple testing. Journal of the Royal statistical society: series B (Methodological). 1995;57(1):289–300.

26. Nadeau C, Bengio Y. Inference for the generalization error. Machine Learning. 2003;52(3):239–81.

27. Bouckaert R, Frank E, Dai H, Srikant R, Zhang C. Evaluating the replicability of significance tests for comparing learning algorithms. Advances in Knowledge Discovery and Data Mining, Proceedings. 2004;3056:3–12.

28. Rahmim A, Salimpour Y, Jain S, Blinder SA, Klyuzhin IS, Smith GS, et al. Application of texture analysis to DAT SPECT imaging: Relationship to clinical assessments. Neuroimage Clin. 2016;12:e1–e9.

29. Rahmim A, Huang P, Shenkov N, Fotouhi S, Davoodi-Bojd E, Lu L, et al. Improved prediction of outcome in Parkinson’s disease using radiomics analysis of longitudinal DAT SPECT images. Neuroimage Clin. 2017;16:539–44.

30. Weintraub D, Claassen DO. Impulse Control and Related Disorders in Parkinson’s Disease. Int Rev Neurobiol. 2017;133:679–717.

31. Jesús S, Periñán MT, Cortés C, Buiza-Rueda D, Macías-García D, Adarmes A, et al. Integrating genetic and clinical data to predict impulse control disorders in Parkinson’s disease. Eur J Neurol. 2021;28(2):459–68.

32. Ricciardi L, Lambert C, De Micco R, Morgante F, Edwards M. Can we predict development of impulsive-compulsive behaviours in Parkinson’s disease? J Neurol Neurosurg Psychiatry. 2018;89(5):476–81.

33. Hinkle JT, Perepezko K, Gonzalez LL, Mills KA, Pontone GM. Apathy and Anxiety in De Novo Parkinson’s Disease Predict the Severity of Motor Complications. Mov Disord Clin Pract. 2021;8(1):76–84.

34. Meng D, Jin Z, Wang Y, Fang B. Longitudinal cognitive changes in patients with early Parkinson’s disease and neuropsychiatric symptoms. CNS Neurosci Ther. 2023;29(8):2259–66.

35. Roussakis AA, Lao-Kaim NP, Piccini P. Brain Imaging and Impulse Control Disorders in Parkinson’s Disease. Curr Neurol Neurosci Rep. 2019;19(9):67.

36. Hernadi G, Perlaki G, Kovacs M, Pinter D, Orsi G, Janszky J, et al. White matter hyperintensities associated with impulse control disorders in Parkinson’s Disease. Sci Rep. 2023;13(1):10594.

37. Gan C, Zhang H, Sun H, Cao X, Wang L, Zhang K, et al. Aberrant brain topological organization and granger causality connectivity in Parkinson’s disease with impulse control disorders. Front Aging Neurosci. 2024;16:1364402.

