## supplemental material S1 for "Prediction of impulse control disorders in Parkinson’s disease: a longitudinal machine learning study"

### SUPPLEMENTARY MATERIAL

#### S1. TRIPOD+AI statement

TRIPOD+AI

Version: 11-January-2024

| Section/Topic | Item | Development / evaluation[1] | Checklist item | Reported on page |
|---|---|---|---|---|
| **TITLE** | | | | |
| *Title* | 1 | D;E | Identify the study as developing or evaluating the performance of a multivariable prediction model, the target population, and the outcome to be predicted | 1 |
| **ABSTRACT** | | | | |
| *Abstract* | 2 | D;E | See TRIPOD+AI for Abstracts checklist | 2 |
| INTRODUCTION | | | | |
| *Background* | 3a | D;E | Explain the healthcare context (including whether diagnostic or prognostic) and rationale for developing or evaluating the prediction model, including references to existing models | 3 |
| | 3b | D;E | Describe the target population and the intended purpose of the prediction model in the context of the care pathway, including its intended users (e.g., healthcare professionals, patients, public) | 3 |
| | 3c | D;E | Describe any known health inequalities between sociodemographic groups | 3 |
| *Objectives* | 4 | D;E | Specify the study objectives, including whether the study describes the development or validation of a prediction model (or both) | 4 |
| METHODS | | | | |
| *Data* | 5a | D;E | Describe the sources of data separately for the development and evaluation datasets (e.g., randomised trial, cohort, routine care or registry data), the rationale for using these data, and representativeness of the data | 4 |
| | 5b | D;E | Specify the dates of the collected participant data, including start and end of participant accrual; and, if applicable, end of follow-up | Sup p. 3 |
| *Participants* | 6a | D;E | Specify key elements of the study setting (e.g., primary care, secondary care, general population) including the number and location of centres | Sup p. 3 |
| | 6b | D;E | Describe the eligibility criteria for study participants | 4 |
| | 6c | D;E | Give details of any treatments received, and how they were handled during model development or evaluation, if relevant | Sup p. 3 |
| *Data preparation* | 7 | D;E | Describe any data pre-processing and quality checking, including whether this was similar across relevant sociodemographic groups | 5-6 |
| *Outcome* | 8a | D;E | Clearly define the outcome that is being predicted and the time horizon, including how and when assessed, the rationale for choosing this outcome, and whether the method of outcome assessment is consistent across sociodemographic groups | 5 |
| | 8b | D;E | If outcome assessment requires subjective interpretation, describe the qualifications and demographic characteristics of the outcome assessors | 5 |
| | 8c | D;E | Report any actions to blind assessment of the outcome to be predicted | n/a |
| *Predictors* | 9a | D | Describe the choice of initial predictors (e.g., literature, previous models, all available predictors) and any pre-selection of predictors before model building | 5-6 |
| | 9b | D;E | Clearly define all predictors, including how and when they were measured (and any actions to blind assessment of predictors for the outcome and other predictors) | 5-6, Sup p. 4-7, 13 |
| | 9c | D;E | If predictor measurement requires subjective interpretation, describe the qualifications and demographic characteristics of the predictor assessors | n/a |
| *Sample size* | 10 | D;E | Explain how the study size was arrived at (separately for development and evaluation), and justify that the study size was sufficient to answer the research question. Include details of any sample size calculation | 7 |
| *Missing data* | 11 | D;E | Describe how missing data were handled. Provide reasons for omitting any data | 6 |
| *Analytical methods* | 12a | D | Describe how the data were used (e.g., for development and evaluation of model performance) in the analysis, including whether the data were partitioned, considering any sample size requirements | 6-7 |
| | 12b | D | Depending on the type of model, describe how predictors were handled in the analyses (functional form, rescaling, transformation, or any standardisation). | 6-7 |
| | 12c | D | Specify the type of model, rationale[2], all model-building steps, including any hyperparameter tuning, and method for internal validation | 6-7 |
| | 12d | D;E | Describe if and how any heterogeneity in estimates of model parameter values and model performance was handled and quantified across clusters (e.g., hospitals, countries). See TRIPOD-Cluster for additional considerations[3] | 6-7 |
| | 12e | D;E | Specify all measures and plots used (and their rationale) to evaluate model performance (e.g., discrimination, calibration, clinical utility) and, if relevant, to compare multiple models | 7 |
| | 12f | E | Describe any model updating (e.g., recalibration) arising from the model evaluation, either overall or for particular sociodemographic groups or settings | 14, Sup p. 14-20 |
| | 12g | E | For model evaluation, describe how the model predictions were calculated (e.g., formula, code, object, application programming interface) | 6-7 |
| *Class imbalance* | 13 | D;E | If class imbalance methods were used, state why and how this was done, and any subsequent methods to recalibrate the model or the model predictions | 6-7 |
| *Fairness* | 14 | D;E | Describe any approaches that were used to address model fairness and their rationale | 6-7 |
| *Model output* | 15 | D | Specify the output of the prediction model (e.g., probabilities, classification). Provide details and rationale for any classification and how the thresholds were identified | 7 |

[1] D=items relevant only to the development of a prediction model; E=items relating solely to the evaluation of a prediction model; D;E=items applicable to both the development and evaluation of a prediction model

[2] Separately for all model building approaches.

[3] TRIPOD-Cluster is a checklist of reporting recommendations for studies developing or validating models that explicitly account for clustering or explore heterogeneity in model performance (eg, at different hospitals or centres). Debray et al, BMJ 2023; 380: e071018 [DOI: 10.1136/bmj-2022-071018]

| | | | | |
|---|---|---|---|---|
| *Training versus evaluation* | 16 | D;E | Identify any differences between the development and evaluation data in healthcare setting, eligibility criteria, outcome, and predictors | 7 |
| *Ethical approval* | 17 | D;E | Name the institutional research board or ethics committee that approved the study and describe the participant-informed consent or the ethics committee waiver of informed consent | 4 |
| **OPEN SCIENCE** | | | | |
| *Funding* | 18a | D;E | Give the source of funding and the role of the funders for the present study | 17-18 |
| *Conflicts of interest* | 18b | D;E | Declare any conflicts of interest and financial disclosures for all authors | 18 |
| *Protocol* | 18c | D;E | Indicate where the study protocol can be accessed or state that a protocol was not prepared | n/a |
| *Registration* | 18d | D;E | Provide registration information for the study, including register name and registration number, or state that the study was not registered | 4 |
| *Data sharing* | 18e | D;E | Provide details of the availability of the study data | 18 |
| *Code sharing* | 18f | D;E | Provide details of the availability of the analytical code[4] | 18 |
| **PATIENT & PUBLIC INVOLVEMENT** | | | | |
| *Patient & Public Involvement* | 19 | D;E | Provide details of any patient and public involvement during the design, conduct, reporting, interpretation, or dissemination of the study or state no involvement. | n/a |
| **RESULTS** | | | | |
| *Participants* | 20a | D;E | Describe the flow of participants through the study, including the number of participants with and without the outcome and, if applicable, a summary of the follow-up time. A diagram may be helpful. | 8-9 |
| | 20b | D;E | Report the characteristics overall and, where applicable, for each data source or setting, including the key dates, key predictors (including demographics), treatments received, sample size, number of outcome events, follow-up time, and amount of missing data. A table may be helpful. Report any differences across key demographic groups. | 10, Sup. p. 8-13 |
| | 20c | E | For model evaluation, show a comparison with the development data of the distribution of important predictors (demographics, predictors, and outcome). | Sup. p. 8-13 |
| *Model development* | 21 | D;E | Specify the number of participants and outcome events in each analysis (e.g., for model development, hyperparameter tuning, model evaluation) | 11 |
| *Model specification* | 22 | D | Provide details of the full prediction model (e.g., formula, code, object, application programming interface) to allow predictions in new individuals and to enable third-party evaluation and implementation, including any restrictions to access or re-use (e.g., freely available, proprietary)[5] | 18 |
| *Model performance* | 23a | D;E | Report model performance estimates with confidence intervals, including for any key subgroups (e.g., sociodemographic). Consider plots to aid presentation. | 11-13, Sup. p. 14-20 |
| | 23b | D;E | If examined, report results of any heterogeneity in model performance across clusters. See TRIPOD Cluster for additional details[3]. | 13 |
| *Model updating* | 24 | E | Report the results from any model updating, including the updated model and subsequent performance | 14, Sup. p. 17-19 |
| **DISCUSSION** | | | | |
| *Interpretation* | 25 | D;E | Give an overall interpretation of the main results, including issues of fairness in the context of the objectives and previous studies | 14-17 |
| *Limitations* | 26 | D;E | Discuss any limitations of the study (such as a non-representative sample, sample size, overfitting, missing data) and their effects on any biases, statistical uncertainty, and generalizability | 17 |
| *Usability of the model in the context of current care* | 27a | D | Describe how poor quality or unavailable input data (e.g., predictor values) should be assessed and handled when implementing the prediction model | n/a |
| | 27b | D | Specify whether users will be required to interact in the handling of the input data or use of the model, and what level of expertise is required of users | n/a |
| | 27c | D;E | Discuss any next steps for future research, with a specific view to applicability and generalizability of the model | 17 |

From: Collins GS, Moons KGM, Dhiman P, et al. *BMJ* 2024;385:e078378. doi:10.1136/bmj-2023-078378

[4] This relates to the analysis code, for example, any data cleaning, feature engineering, model building, evaluation.
[5] This relates to the code to implement the model to get estimates of risk for a new individual.

#### S2. AmsterdamUMC dataset

Baseline data was collected for routine clinical care and follow-up data was collected across four separate longitudinal studies:

1) Amsterdam 1: this retrospective study (previously described in (1)) collected data between May 2008 and December 2011. Briefly, the original cohort consisted of 31 *de novo* PD patients that underwent DAT-SPECT imaging, subsequently initiated dopaminergic therapy and were assessed on development of ICD approximately three years later through questionnaires and phone interviews.
2) Amsterdam 2: this retrospective study collected data between May 2008 and July 2012. The original cohort consisted of 116 participants that filled out neuropsychiatric symptom questionnaires and information on medication use and quality of life at on average 3.8 years after diagnosis.
3) Amsterdam 3: this prospective study (previously described in (2)) collected baseline data between February 2010 and November 2012. The original cohort consisted of 20 non-demented individuals with PD that underwent extensive clinical assessment at time of diagnosis and approximately three years later.
4) Amsterdam 4: this prospective study was initiated to study the predictors and development of ICDs in PD, with baseline data collection between August 2014 and October 2018. The original sub-cohort consisted of 31 *de novo* PD patients that underwent DAT-SPECT imaging at baseline, and additional extensive clinical assessment at baseline, six-months, one year, two years and four years follow-up.

#### S3. Harmonization of clinical features between cohorts

**Table S3** Measurement of clinical features and steps to harmonize between cohorts.

| Outcome | PPMI measure | AMSTERDAMUMC measure | Harmonization step |
|---|---|---|---|
| Disease stage | Hoehn & Yahr stage | Modified Hoehn & Yahr stage | We recoded stages 1.5 and 2.5 (from the modified version) to stage 2 (3) |
| PD motor symptom severity | Movement Disorder Society-Unified PD Rating Scale motor score (MDS-UPDRS) | UPDRS part III motor score | We added 7 points to the original UPDRS-III total score (4). |
| Depressive symptoms | Geriatric Depression Scale-15 | Beck Depression Inventory | We recoded total scores into four severity classifications (None, Mild, Moderate, Severe) (5, 6) |
| Anxiety | Trait subscale of the State-Trait Anxiety Inventory | Beck Anxiety Inventory | We recoded total scores into three severity classifications (None/Minimal, Moderate, Severe/High) (7, 8). |
| Apathy | MDS-UPDRS part I item 5 | UPDRS part I item 4 | None |
| Global cognitive function | Montreal Cognitive Assessment (MoCA) | Mini-Mental State Examination (MMSE) | We converted the MoCA scores to an MMSE score (9) |

#### S4. DaT-SPECT image acquisition, reconstruction and post-acquisition processing

In the PPMI sample, [$^{123}$I]FP-CIT SPECT was used to measure presynaptic striatal dopamine transporter density. Patients were pre-treated with stable iodine to reduce the thyroid uptake of [$^{123}$I]FP-CIT. Four hours (± 30 minutes) before image acquisition, $^{123}$I-DaTscan™ was intravenously administered in a dose between 111-185 mBq. Raw projection data was acquired into a 128x128 matrix with low-energy high resolution (LEHR) parallel hole collimators using a circular step and shoot mode with each step moving 3 degrees for a total of 60 projections per detector (180 degrees rotation per detector) for dual head systems, 30 seconds per projection, with a 20 % energy window centered on 159 keV (±10 %) for 123IDaTscan™ and acquisition zoom of 1.23.

SPECT raw projection data was imported to a HERMES (Hermes Medical Solutions, Skeppsbron 44, 111 30 Stockholm, Sweden) system for iterative (HOSEM) reconstruction. The HOSEM reconstructed files were then transferred to the PMOD (PMOD Technologies, Zurich, Switzerland) where Attenuation correction ellipses where drawn on the images and a Chang 0 attenuation correction was applied images utilizing a site specific mu that was empirically derived from phantom data acquired during site initiation for the trial. Once attenuation correction was completed a standard Gaussian 3D 6.0 mm filter was applied.

In the AmsterdamUMC sample, [$^{123}$I]FP-CIT SPECT was used to measure presynaptic striatal dopamine transporter density. Patients orally received potassium perchlorate to block thyroid update of free radioactive iodide. Three hours before image acquisition, [$^{123}$I]FP-CIT was intravenously administered in a dose of approximately 185 MBq (specific activity >185MBq/nmol; radiochemical purity >99%; produced according to good manufacturing practices criteria at GE Healthcare, Eindhoven, The Netherlands). Static images were subsequently obtained for 30 minutes (60×30 s views per head over a 180° orbit on a 128×128-pixel matrix) using a dual-head gamma camera (E.Cam; Siemens, Munich, Germany) with a fan-beam collimator. Image reconstruction was performed using a filtered back projection with a Butterworth filter (order 8, cut-off 0.6 cycles/cm).

#### S5. Feature extraction

##### Radiomic features

Recent studies have utilized morphological and advanced intensity histogram-based features, termed radiomics, for investigating the utility of DAT-SPECT beyond the standard task of confirmation of initial PD diagnosis. Radiomic features, that constitute a more detailed representation of the imaging properties, are more sensitively associated with motor and cognitive decline scores compared to the average signal intensity of striatal regions, given SBRs (10). Of note, it was also shown to hold potential in predicting future motor symptom severity (11). We used the Pyradiomics library (12) to extract radiomic features from each striatal VOI, considering the original as well as filtered version of the images using Wavelet decompositions and Laplacian of Gaussian (LoG) filters.

We used a fixed bin-width option for image discretization prior to feature extraction, to maintain the relative difference between the discretized image contrasts, as significantly lower DAT striatal uptake was previously reported in individuals with an ICD (13, 14). We considered the average intensity ranges of each region in this process, to ensure that the number of bins of the discretized VOIs ranged between 16 and 130, as recommended by the pyradiomics developers. We extracted 19 first order statistics, 24 features from the Gray Level Co-occurrence Matrix (GLCM), 16 features from the Gray Level Run Length Matrix (GLRLM), 16 features from the Gray Level Size Zone (GLSZM), 5 features from the Neighbouring Gray Tone Difference Matrix (NGTDM) and 14 features from the Gray Level Dependence Matrix (GLDM), from original image, as well as from 8 different wavelet decompositions and 5 different LoG sigma levels (1-5 mm), resulting into a total of 1232 features per VOI.

##### Latent features

We implemented a 3D deep autoencoder in Tensorflow/Keras 2.0 DL python library (https://keras.io/), by adjusting the architecture previously described in (15), to extract latent from the striatal regions. The autoencoder network was trained on cropped volumes defined with a fixed-size bounding box of 32x32x24 voxels applied to the segmented striatum on the preprocessed scans. Image intensities of the cropped volumes were further normalized to [0-1] range. The DAT-SCANS of the healthy controls were also included to increase the sample size. Artificial image augmentation was also employed by horizontal flipping (left<->right) of a random fraction of the input images within each training batch. The network's encoder part consisted of three consecutive blocks, each including a 3 × 3 x 3 convolutional layer with a leaky rectified linear unit (Leaky ReLU), followed by a batch normalization layer, and a 2 × 2 x 2 max-pooling layer. The decoder part of the network consisted of three consecutive blocks, including a batch normalization layer, a 3 × 3 x 3 convolutional layer with ReLU activation and 2 × 2 x 2 upsampling layers. A final 3D convolution layer without activation was used on top. A fully connected layer with 1024 nodes was used in the network bottleneck. Model training considered Adam optimization and the Mean Squared Error loss. The trained model bottleneck was used to extract 1024 latent features from the PD images for the subsequent models.

#### S6. Demographic and clinical data of the PPMI and AmsterdamUMC datasets

**Table S6** Demographic and clinical characteristics of the PPMI and AmsterdamUMC datasets

| | **AmsterdamUMC (n=67)** | **PPMI (n=311)** | **P-value** |
|---|---|---|---|
| **Age (years)** | 63.0 (±11.2) | 61.2 (±9.6) | 0.16 |
| **Sex (% female)** | 22 (32.8%) | 107 (34.4%) | 0.89 |
| **Education (years)** | 15.2 (±4.7) | 15.6 (±2.9) | 0.99 |
| **Duration of follow-up (years)** | 3.3 (±1.3) | 8.1 (±3.0) | < 0.0001 |
| **Age at symptom onset (years)** | 60.0 (±10.8) | 59.2 (±9.8) | 0.7 |
| **MDS-UPDRS part III** | 28.7 (±10.7) | 21.0 (±9.1) | < 0.0001 |
| **Hoehn & Yahr stage** | | | < 0.0001 |
| **1** | 12 (17.9%) | 137 (44.1%) | |
| **2** | 51 (76.1%) | 172 (55.3%) | |
| **3** | 4 (6.0%) | 2 (0.6%) | |
| **MMSE** | 28.3 (±1.7) | 29.2 (±1.2) | < 0.0001 |
| **Depressive symptom severity** | | | 0.0001 |
| **Minimal** | 46 (68.7%) | 277 (89.1%) | |
| **Mild** | 17 (25.4%) | 23 (7.4%) | |
| **Moderate** | 4 (6.0%) | 8 (2.6%) | |
| **Severe** | 0 (0.0%) | 3 (1.0%) | |
| **Anxiety symptom severity** | | | < 0.0001 |
| **No to minimal** | 16 (23.9%) | 243 (78.1%) | |
| **Moderate** | 22 (32.8%) | 62 (19.9%) | |
| **Severe** | 29 (43.3%) | 6 (1.9%) | |
| **(MDS-)UPDRS part I Apathy score** | | | 0.004 |
| **0** | 48 (71.6%) | 265 (85.2%) | |
| **1** | 13 (19.4%) | 40 (12.9%) | |
| **2** | 4 (6.0%) | 5 (1.6%) | |
| **3** | 2 (3.0%) | 0 (0.0%) | |
| **4** | 0 (0.0%) | 1 (0.3%) | |
| **Psychotropic medication use** | | | 0.11 |
| **SSRI** | 5 (7.5%) | 30 (9.6%) | |
| **SSRI + Benzodiazepines** | 1 (1.5%) | 0 (0.0%) | |
| **Benzodiazepines** | 2 (3.0%) | 30 (9.6%) | |
| **None** | 59 (88.1%) | 251 (80.7%) | |
| **ICD group** | | | 0.079 |
| **No ICD** | 42 (62.7%) | 157 (50.5%) | |
| **ICB** | 5 (7.5%) | 53 (17.0%) | |
| **ICD** | 20 (29.9%) | 101 (32.5%) | |
| **Time to ICD/ICB (years)** | 3.2 (±1.2) | 4.3 (±2.6) | 0.13 |

*Abbreviations:* MMSE = Mini-mental state examination; MDS-UPDRS = Movement Disorders Society – Unified PD Rating Scale

**Table S7** Group statistics of the ICD, No ICD groups in the full combined dataset (PPMI+AmsterdamUMC).

| | No ICD (n=199) | ICB (n=58) | ICD (n=121) |
|---|---|---|---|
| **Age (years)** | 62.8 (±9.7) | 62.9 (±9.0) | 58.7 (±10.2) |
| **Sex (% female)** | 77 (38.7%) | 15 (25.9%) | 37 (30.6%) |
| **Education (years)** | 15.7 (±3.1) | 15.9 (±3.0) | 15.3 (±3.0) |
| **Duration of follow-up (years)** | 6.6 (±3.4) | 7.9 (±3.2) | 8.1 (±3.1) |
| **Age at symptom onset (years)** | 60.8 (±9.7) | 60.6 (±9.0) | 56.5 (±10.2) |
| **(MDS-)UPDRS part III** | 22.5 (±10.0) | 22.6 (±7.8) | 22.2 (±10.5) |
| **Hoehn & Yahr stage** | | | |
| **1** | 77 (38.7%) | 15 (25.9%) | 57 (47.1%) |
| **2** | 118 (59.3%) | 42 (72.4%) | 63 (52.1%) |
| **3** | 4 (2.0%) | 1 (1.7%) | 1 (0.8%) |
| **LEDD total (at follow-up)** | 683.9 (±512.3) | 595.4 (±483.0) | 549.1 (±416.8) |
| **LEDD levodopa** | 521.1 (±461.5) | 433.4 (±439.7) | 350.4 (±404.3) |
| **LEDD dopamine agonists** | 62.0 (±118.5) | 85.5 (±129.1) | 105.7 (±124.2) |
| **LEDD max. (across follow-ups)** | 774.4 (±561.5) | 650.6 (±543.7) | 564.9 (±432.2) |
| **LEDD levodopa max.** | 592.0 (±517.3) | 484.4 (±514.7) | 346.6 (±413.0) |
| **LEDD dopamine agonists max.** | 98.2 (±141.2) | 112.7 (±153.7) | 134.4 (±143.7) |
| **MMSE** | 29.0 (±1.3) | 28.9 (±1.4) | 29.1 (±1.3) |
| **Depressive symptom severity** | | | |
| **Minimal** | 172 (86.4%) | 54 (93.1%) | 97 (80.2%) |
| **Mild** | 19 (9.5%) | 4 (6.9%) | 17 (14.0%) |
| **Moderate** | 6 (3.0%) | 0 (0.0%) | 6 (5.0%) |
| **Severe** | 2 (1.0%) | 0 (0.0%) | 1 (0.8%) |
| **Anxiety symptom severity** | | | |
| **No to minimal** | 142 (71.4%) | 43 (74.1%) | 74 (61.2%) |
| **Moderate** | 40 (20.1%) | 14 (24.1%) | 30 (24.8%) |
| **Severe** | 17 (8.5%) | 1 (1.7%) | 17 (14.0%) |
| **(MDS-)UPDRS part I Apathy score** | | | |
| **0** | 173 (86.9%) | 47 (81.0%) | 93 (76.9%) |
| **1** | 20 (10.1%) | 11 (19.0%) | 22 (18.2%) |
| **2** | 3 (1.5%) | 0 (0.0%) | 6 (5.0%) |
| **3** | 2 (1.0%) | 0 (0.0%) | 0 (0.0%) |
| **4** | 1 (0.5%) | 0 (0.0%) | 0 (0.0%) |

Abbreviations: LEDD = Levodopa Equivalent Daily Dosage; (MDS-)UPDRS = (Movement Disorders Society version) Unified Parkinson's Disease Rating Scale; MMSE = Mini-Mental State Examination

#### S8. Univariate statistics of the clinical features for comparison of ICD versus No ICD

**Table S8** Statistics of the univariate group comparisons between the ICD and No ICD group

| Outcome | Model | B | SE | 95CI- | 95CI+ | P-value |
|---|---|---|---|---|---|---|
| **Sex[2]** | crude | OR = 1.431 | | 0.864 | 2.394 | 0.150 |
| | AmsterdamUMC | OR = 0.929 | | 0.268 | 3.406 | 1.000 |
| | PPMI | OR = 1.583 | | 0.903 | 2.813 | 0.111 |
| **Education[1,*]** | crude | -0.418 | 0.379 | -1.165 | 0.328 | 0.271 |
| | Corrected (Age + Sex) | -0.496 | 0.389 | -1.262 | 0.270 | 0.203 |
| | Group * PPMI (corrected for Age + Sex) | -0.169 | 0.391 | -0.939 | 0.601 | 0.666 |
| **Age at symptom onset[1]** | **crude** | **-4.285** | **1.161** | **-6.570** | **-2.001** | **<0.001** |
| | **Corrected (Sex)** | **-4.362** | **1.163** | **-6.651** | **-2.074** | **<0.001** |
| | Group * AmsterdamUMC (corrected for Age + Sex) | -2.226 | 2.809 | -7.754 | 3.302 | 0.429 |
| | **Group * PPMI (corrected for Age + Sex)** | **-4.807** | **1.280** | **-7.325** | **-2.289** | **<0.001** |
| **MDS-UPDRS part III[1]** | crude | -0.280 | 1.175 | -2.593 | 2.032 | 0.812 |
| | Corrected (Age + Sex) | 0.564 | 1.127 | -1.653 | 2.780 | 0.617 |
| | **Group *** AmsterdamUMC **(corrected for Age + Sex)** | **6.246** | **2.569** | **1.191** | **11.300** | **0.016** |
| | Group * PPMI (corrected for Age + Sex) | -0.747 | 1.239 | -3.184 | 1.690 | 0.547 |
| **Hoehn & Yahr stage[3]** | crude | | | | | 0.311 |
| | AmsterdamUMC | | | | | 0.124 |
| | PPMI | | | | | 0.054 |
| **LEDD[1]** | **crude** | **-134.839** | **56.382** | **-245.782** | **-23.895** | **0.017** |
| | **Corrected (Age + Sex)** | **-195.961** | **56.657** | **-307.451** | **-84.471** | **0.001** |
| | Group * AmsterdamUMC (corrected for Age + Sex) | -37.561 | 146.505 | -325.858 | 250.736 | 0.798 |
| | **Group * PPMI (corrected for Age + Sex)** | **-222.682** | **61.038** | **-342.794** | **-102.569** | **<0.001** |
| **Levodopa use (LEDD)** | **crude** | **-170.6879** | **51.61235** | **-272.24401** | **-69.13179** | **0.00105** |
| | **Corrected (Age + Sex)** | **-197.53495** | **52.17502** | **-300.20217** | **-94.86773** | **0.00018** |
| | Group * AmsterdamUMC (corrected for Age + Sex) | -35.97004 | 129.5969 | -290.98725 | 219.04716 | 0.78154 |
| | **Group * PPMI (corrected for Age + Sex)** | **-227.84173** | **56.65791** | **-339.33161** | **-116.35185** | **<0.0001** |
| **Dopamine agonists use (LEDD)** | **Crude** | **W = 8806.5** | **r = -0.2165** | | | **0.0001** |
| | **PPMI** | **W = 5920** | **r = -0.2403** | | | **0.0001** |
| | AmsterdamUMC | W = 271.5 | r = -0.1198 | | | 0.3453 |
| **MMSE[4]** | Crude | W = 11690 | r = -0.026 | | | 0.637 |
| | PPMI | W = 7157.5 | r = -0.091 | | | 0.143 |
| | **AmsterdamUMC** | **W = 553.5** | **r = -0.261** | | | **0.040** |
| **Depression[3]** | crude | | | | | 0.427 |
| | **AmsterdamUMC** | | | | | **0.039** |
| | PPMI | | | | | 0.850 |
| **Anxiety[3]** | crude | | | | | 0.128 |
| | **AmsterdamUMC** | | | | | **0.007** |
| | PPMI | | | | | 0.046 |

| | | |
|---|---|---|
| **(MDS-)UPDRS part I Apathy score[3]** | **crude** | **0.027** |
| | **AmsterdamUMC** | **0.003** |
| | PPMI | 0.309 |

1 = linear regression analysis, 2 = logistic regression analysis, 3 = Fisher's exact test, 4 = Mann Whitney U test. *group by AmsterdamUMC cohort interaction not shown due to low N with available data in in AmsterdamUMC cohort (n=15).

#### S9. Statistics of the SBRs

**Table S9** Statistics of the specific binding ratios per subgroup

| | No ICD (n=199) | ICB (n=58) | ICD (n=121) |
|---|---|---|---|
| **Left ventral striatum** | 2.0 (±0.5) | 1.9 (±0.5) | 2.0 (±0.5) |
| **Right ventral striatum** | 1.9 (±0.5) | 1.8 (±0.5) | 1.9 (±0.4) |
| **Left posterior putamen** | 1.0 (±0.3) | 0.9 (±0.3) | 1.0 (±0.3) |
| **Right posterior putamen** | 1.0 (±0.3) | 1.1 (±0.3) | 1.1 (±0.3) |
| **Left anterior putamen** | 2.2 (±0.6) | 2.2 (±0.7) | 2.2 (±0.6) |
| **Right anterior putamen** | 2.2 (±0.6) | 2.2 (±0.6) | 2.3 (±0.6) |
| **Left caudate nucleus** | 1.3 (±0.4) | 1.3 (±0.4) | 1.3 (±0.3) |
| **Right caudate nucleus** | 1.3 (±0.4) | 1.2 (±0.3) | 1.3 (±0.3) |
| **Left thalamus** | 0.4 (±0.2) | 0.4 (±0.2) | 0.4 (±0.1) |
| **Right thalamus** | 0.4 (±0.2) | 0.4 (±0.2) | 0.4 (±0.1) |

#### S10. Univariate statistics of the SBRs for comparison of ICD versus No ICD

**Table S10** Statistics of the univariate group comparisons between the ICD and No ICD group for specific binding ratios of striatal ROIs.

| Outcome | Model | B | SE | 95CI- | 95CI+ | P-value | P-value$^{FDR}$ |
|---|---|---|---|---|---|---|---|
| **Left ventral striatum** | crude | 0.0480 | 0.0563 | -0.0628 | 0.1587 | 0.3948 | 1.000 |
| | corrected | -0.0245 | 0.0549 | -0.1324 | 0.0834 | 0.6554 | 1.000 |
| | Group * AmsterdamUMC (corrected for Age + Sex) | -0.0935 | 0.1262 | -0.3419 | 0.1548 | 0.4592 | 1.000 |
| | Group * PPMI (corrected for Age + Sex) | -0.0086 | 0.0609 | -0.1283 | 0.1111 | 0.8879 | 1.000 |
| **Right ventral striatum** | crude | 0.0342 | 0.0532 | -0.0705 | 0.1389 | 0.5207 | 1.000 |
| | corrected | -0.0316 | 0.0520 | -0.1339 | 0.0707 | 0.5436 | 1.000 |
| | Group * AmsterdamUMC (corrected for Age + Sex) | -0.1743 | 0.1193 | -0.4091 | 0.0605 | 0.1451 | 0.870 |
| | Group * PPMI (corrected for Age + Sex) | 0.0013 | 0.0575 | -0.1119 | 0.1145 | 0.9818 | 1.000 |
| **Left posterior putamen** | crude | 0.0040 | 0.0338 | -0.0626 | 0.0705 | 0.9069 | 1.000 |
| | corrected | 0.0020 | 0.0347 | -0.0663 | 0.0702 | 0.9552 | 1.000 |
| | Group * AmsterdamUMC (corrected for Age + Sex) | 0.0146 | 0.0799 | -0.1425 | 0.1717 | 0.8549 | 1.000 |
| | Group * PPMI (corrected for Age + Sex) | -0.0010 | 0.0385 | -0.0767 | 0.0748 | 0.9799 | 1.000 |
| **Right posterior putamen** | crude | 0.0051 | 0.0362 | -0.0661 | 0.0764 | 0.8879 | 1.000 |
| | corrected | 0.0027 | 0.0370 | -0.0701 | 0.0755 | 0.9419 | 1.000 |
| | Group * AmsterdamUMC (corrected for Age + Sex) | -0.1320 | 0.0848 | -0.2988 | 0.0349 | 0.1207 | 0.845 |
| | Group * PPMI (corrected for Age + Sex) | 0.0338 | 0.0409 | -0.0467 | 0.1142 | 0.4095 | 1.000 |
| **Left anterior putamen** | crude | 0.0345 | 0.0715 | -0.1062 | 0.1751 | 0.6298 | 1.000 |
| | corrected | 0.0008 | 0.0731 | -0.1429 | 0.1446 | 0.9908 | 1.000 |
| | Group * AmsterdamUMC (corrected for Age + Sex) | -0.0386 | 0.1681 | -0.3694 | 0.2922 | 0.8186 | 1.000 |
| | Group * PPMI (corrected for Age + Sex) | 0.0099 | 0.0811 | -0.1496 | 0.1694 | 0.9025 | 1.000 |
| **Right anterior putamen** | crude | 0.0675 | 0.0707 | -0.0717 | 0.2066 | 0.3407 | 1.000 |
| | corrected | 0.0380 | 0.0725 | -0.1046 | 0.1807 | 0.6001 | 1.000 |
| | Group * AmsterdamUMC (corrected for Age + Sex) | -0.2086 | 0.1662 | -0.5355 | 0.1183 | 0.2102 | 1.000 |
| | Group * PPMI (corrected for Age + Sex) | 0.0949 | 0.0801 | -0.0627 | 0.2526 | 0.2368 | 1.000 |
| **Left caudate nucleus** | crude | 0.0438 | 0.0411 | -0.0371 | 0.1246 | 0.2880 | 1.000 |
| | corrected | -0.0044 | 0.0394 | -0.0819 | 0.0730 | 0.9105 | 1.000 |
| | Group * AmsterdamUMC (corrected for Age + Sex) | -0.0824 | 0.0905 | -0.2604 | 0.0956 | 0.3632 | 1.000 |
| | Group * PPMI (corrected for Age + Sex) | 0.0136 | 0.0436 | -0.0723 | 0.0994 | 0.7562 | 1.000 |
| **Right caudate nucleus** | crude | 0.0451 | 0.0398 | -0.0331 | 0.1233 | 0.2574 | 1.000 |
| | corrected | -0.0056 | 0.0380 | -0.0804 | 0.0692 | 0.8831 | 1.000 |
| | **Group * AmsterdamUMC (corrected for Age + Sex)** | **-0.1743** | **0.0868** | **-0.3452** | **-0.0035** | **0.0455** | 0.364 |
| | Group * PPMI (corrected for Age + Sex) | 0.0333 | 0.0419 | -0.0490 | 0.1157 | 0.4265 | 1.000 |

Negative B-values represent lower binding ratios in the ICD group compared with No ICD.
Abbreviations: AmsterdamUMC – Amsterdam cohort; FDR – False discovery rate; ICB – Impulse control behaviors; ICD – Impulse control disorder; PPMI – Parkinson's Progression Markers Initiative cohort; ROI – Region of interest.

#### S11. Statistics of the SNPs

**Table S11** Statistics of the univariate group comparisons between the ICD and No ICD group for SNPs.

| Outcome | rs ID | P-value | P-value$^{FDR}$ |
|---|---|---|---|
| COMT | rs4680 | 0.7242 | 1.0000 |
| DDC | rs3837091 | 0.4027 | 1.0000 |
| DDC | rs1451375 | 0.7027 | 1.0000 |
| DRD1 | rs5326 | 0.2218 | 1.0000 |
| **DRD2/ANKK1** | rs1800497 | **0.0279** | **0.4460** |
| DRD3 | rs6280 | 0.6413 | 1.0000 |
| GRIN2B | rs7301328 | 0.8698 | 1.0000 |
| HTR2A | rs6313 | 0.9339 | 1.0000 |
| NOS1 | rs2682826 | 0.2439 | 1.0000 |
| NRG1 | rs3924999 | 0.8191 | 1.0000 |
| OPRK1 | rs702764 | 0.4324 | 1.0000 |
| OPRM1 | rs1799971 | 0.3224 | 1.0000 |
| OPRM1 | rs677830 | 0.7152 | 1.0000 |
| SLC22A1 | rs628031 | 0.5124 | 1.0000 |
| TPH2_a | rs4290270 | 0.3009 | 1.0000 |
| TPH2_b | rs7305115 | 0.3621 | 1.0000 |

Fisher's exact tests. Abbreviations: FDR – False discovery rate; ICD – Impulse control disorder; SNP – single nucleotide polymorphisms.

#### S12. Post-hoc machine learning analyses:

##### A. Comparison ICD versus No ICD with cut-off at 4th year

As the prediction of an event further into the future may be more difficult, we performed an exploratory analysis using time to ICD within 4 years. Here, we classified patients only as ICD if they had developed ICD *within 4 years* after baseline. Patients that developed ICD after 4 years were removed from these analyses. We contrasted this subgroup with patients that did not show any positive screen for ICD for more than 7 years after baseline. This led to the exclusion of the AmsterdamUMC cohorts for the No ICD group. The characteristics and univariate group comparisons of this subsample are reported in the Tables S12-14.

**Table S12** Demographic and clinical characteristics of the PPMI and AmsterdamUMC cohorts with ICD development within 4 years or no ICD developed after >4 years.

| | AmsterdamUMC (n=17) | PPMI (n=159) |
|---|---|---|
| **Age (years)** | 61.8 (±9.7) | 60.3 (±9.0) |
| **Sex (% female)** | 6 (35.3%) | 63 (39.6%) |
| **Education (years)** | 10.1 (±4.1) | 15.9 (±2.8) |
| **Duration of follow-up (years)** | 3.2 (±0.9) | 9.2 (±2.2) |
| **Age at symptom onset (years)** | 58.7 (±9.9) | 58.5 (±9.4) |
| **MDS-UPDRS part III** | 32.4 (±10.3) | 20.0 (±9.4) |
| **Hoehn & Yahr stage** | | |
| **1** | 1 (5.9%) | 77 (48.4%) |
| **2** | 15 (88.2%) | 81 (50.9%) |
| **3** | 1 (5.9%) | 1 (0.6%) |
| **MMSE** | 27.7 (±1.6) | 29.2 (±1.2) |
| **Depressive symptom severity** | | |
| **Minimal** | 9 (52.9%) | 138 (86.8%) |
| **Mild** | 6 (35.3%) | 16 (10.1%) |
| **Moderate** | 2 (11.8%) | 3 (1.9%) |
| **Severe** | 0 (0.0%) | 2 (1.3%) |
| **Anxiety symptom severity** | | |
| **No to minimal** | 2 (11.8%) | 122 (76.7%) |
| **Moderate** | 3 (17.6%) | 33 (20.8%) |
| **Severe** | 12 (70.6%) | 4 (2.5%) |
| **(MDS-)UPDRS part I Apathy score** | | |
| **0** | 9 (52.9%) | 136 (85.5%) |
| **1** | 5 (29.4%) | 21 (13.2%) |
| **2** | 3 (17.6%) | 2 (1.3%) |
| **No ICD / ICD** | 0 (0.0%) / 17 (100.0%) | 95 (59.7%) / 64 (40.3%) |
| **Time to ICD/ICB (years)** | 3.0 (±1.1) | 2.5 (±1.2) |

*Abbreviations:* MMSE = Mini-mental state examination; MDS-UPDRS = Movement Disorders Society – Unified PD Rating Scale

**Table S13** Group statistics of the ICD and No ICD groups with ICD development within 4 years or no ICD developed after >4 years.

| | No ICD (n=95) | ICD (n=81) |
|---|---|---|
| **Age (years)** | 61.5 (±8.1) | 59.3 (±10.0) |
| **Sex (% female)** | 43 (45.3%) | 26 (32.1%) |
| **Education (years)** | 16.2 (±2.8) | 15.1 (±3.0) |
| **Duration of follow-up (years)** | 9.8 (±1.4) | 7.3 (±3.3) |
| **Age at symptom onset (years)** | 59.7 (±8.6) | 57.0 (±10.1) |
| **(MDS-)UPDRS part III** | 19.4 (±9.1) | 23.4 (±10.9) |
| **Hoehn & Yahr stage** | | |
| **1** | 45 (47.4%) | 33 (40.7%) |
| **2** | 49 (51.6%) | 47 (58.0%) |
| **3** | 1 (1.1%) | 1 (1.2%) |
| **LEDD** | 874.4 (±570.9) | 415.8 (±290.0) |
| **MMSE** | 29.2 (±1.3) | 29.0 (±1.4) |
| **Depressive symptom severity** | | |
| **Minimal** | 86 (90.5%) | 61 (75.3%) |
| **Mild** | 7 (7.4%) | 15 (18.5%) |
| **Moderate** | 1 (1.1%) | 4 (4.9%) |
| **Severe** | 1 (1.1%) | 1 (1.2%) |
| **Anxiety symptom severity** | | |
| **No to minimal** | 82 (86.3%) | 42 (51.9%) |
| **Moderate** | 11 (11.6%) | 25 (30.9%) |
| **Severe** | 2 (2.1%) | 14 (17.3%) |
| **(MDS-)UPDRS part I Apathy score** | | |
| **0** | 86 (90.5%) | 59 (72.8%) |
| **1** | 8 (8.4%) | 18 (22.2%) |
| **2** | 1 (1.1%) | 4 (4.9%) |

Abbreviations: LEDD = Levodopa Equivalent Daily Dosage; (MDS-)UPDRS = (Movement Disorders Society version) Unified Parkinson’s Disease Rating Scale; MMSE = Mini-Mental State Examination

**Table S14** tatistics of the univariate ICD and No ICD group comparisons with ICD development within 4 years or no ICD developed after >4 years

| Outcome | Model | B | SE | 95CI- | 95CI+ | P-value |
|---|---|---|---|---|---|---|
| **Sex[2]** | crude | OR = 1.7437 | | 0.9036 | 3.4066 | 0.0890 |
| **Education[1]** | **crude** | **-1.0287** | **0.4585** | **-1.9341** | **-0.1233** | **0.0262** |
| | Corrected (Age + Sex) | -0.8277 | 0.4590 | -1.7343 | 0.0788 | 0.0732 |
| **Age at symptom onset[1]** | crude | -2.6932 | 1.4240 | -5.5042 | 0.1178 | 0.0603 |
| | **Corrected (Sex)** | **-3.6644** | **1.4927** | **-6.6114** | **-0.7175** | **0.0151** |
| **(MDS-)UPDRS part III[1]** | **crude** | **4.0513** | **1.5039** | **1.0831** | **7.0195** | **0.0078** |
| | Corrected (Age + Sex) | 2.1098 | 1.5538 | -0.9573 | 5.1769 | 0.1763 |
| **Hoehn & Yahr Stage[3]** | crude | | | | | 0.7227 |
| **LEDD[1]** | **crude** | **-458.5834** | **71.1792** | **-599.0865** | **-318.0803** | **<0.0001** |
| | **Corrected (Age + Sex)** | **-512.1252** | **76.6616** | **-663.4695** | **-360.7810** | **<0.0001** |
| **MMSE[4]** | Crude | W = 4113 | r = -0.0647 | | | 0.3908 |
| **Depression[3]** | **crude** | | | | | **0.0271** |
| **Anxiety[3]** | **crude** | | | | | **<0.0001** |
| **(MDS-)UPDRS part I Apathy score[3]** | **crude** | | | | | **0.0057** |

1 = linear regression analysis, 2 = logistic regression analysis, 3 = Fisher's exact test, 4 = Mann Whitney U test.
Note that the group x cohort interactions are not provided as all Amsterdam patients screened positive for ICD in this subsample.

Results of the ML analysis are shown in Table S15. All models achieved significantly higher performances compared to the models of the pre-registered analysis. The LR and MLP classifiers presented the highest AUC=0.74±0.06, in the models including only clinical features, with the severity of anxiety at baseline showing the highest importance (MDA=0.17±0.06). Similar to the results of the pre-registered analysis, the additional use of radiomic or deep features did not improve predictability. In the RF model, which presented the highest performance in the Clinical+Radiomics features subset (AUC=0.73±0.07), Anxiety severity and one radiomic feature were found the most important, with MDA=0.04±0.05, and MDA=0.04±0.04, respectively.

**Table S15 Cross-validated performance evaluation of ICD vs non-ICD classification models in the combined dataset (PPMI+AmsterdamUMC) with 4th year prediction cut-off**

| Feature subset | Model | Accuracy | Balanced accuracy | Sensitivity | Specificity | AUC |
|---|---|---|---|---|---|---|
| **Clinical features** | RF | 0.67 ± 0.05 | 0.66 ± 0.05 | 0.62 ± 0.11 | 0.71 ± 0.11 | 0.72 ± 0.05 |
| | LR | 0.70 ± 0.08 | 0.69 ± 0.07 | 0.61 ± 0.12 | 0.78 ± 0.14 | 0.74 ± 0.07 |
| | GBC | 0.62 ± 0.08 | 0.62 ± 0.08 | 0.57 ± 0.11 | 0.67 ± 0.11 | 0.66 ± 0.08 |
| | MLP | 0.71 ± 0.07 | 0.70 ± 0.06 | 0.56 ± 0.10 | 0.83 ± 0.11 | 0.74 ± 0.06 |
| **Clinical + Radiomics features** | RF | 0.66 ± 0.07 | 0.66 ± 0.07 | 0.59 ± 0.11 | 0.72 ± 0.10 | 0.73 ± 0.07 |
| | LR | 0.66 ± 0.07 | 0.66 ± 0.07 | 0.62 ± 0.09 | 0.70 ± 0.11 | 0.72± 0.07 |
| | GBC | 0.66 ± 0.07 | 0.65 ± 0.07 | 0.62 ± 0.12 | 0.69 ± 0.11 | 0.70 ± 0.07 |
| | MLP | 0. 59 ± 0.08 | 0.58 ± 0.07 | 0.47 ± 0.20 | 0.70 ± 0.23 | 0.61 ± 0.07 |
| **Clinical + Latent features*** | RF | 0.62 ± 0.07 | 0.61 ± 0.07 | 0.59 ± 0.12 | 0.64 ± 0.11 | 0.66 ± 0.07 |
| | LR | 0.63 ± 0.08 | 0.62 ± 0.08 | 0.58 ± 0.11 | 0.67 ± 0.11 | 0.66± 0.08 |
| | GBC | 0.59 ± 0.07 | 0.58 ± 0.07 | 0.56 ± 0.12 | 0.61 ± 0.11 | 0.60 ± 0.07 |
| | MLP | 0.63 ± 0.08 | 0.62 ± 0.07 | 0.56 ± 0.12 | 0.69 ± 0.15 | 0.65 ± 0.07 |

Abbreviations: AUC = Area Under Curve, RF = Random Forest, LR = Logistic Regression, GB = Gradient Boosting, MLP = Multi-Layer Perceptron. *Latent features were extracted with the 3D autoencoder network.

We also repeated the same analysis in the PPMI sample alone, to evaluate the contribution of the genetic features (Table S16). In the best performing clinical RF model, Anxiety severity had MDA=0.11±0.07, followed by REM-sleep behavioral disorder MDA=0.04±0.05. Overall, genetic variables did not significantly improve the models. Of note, we observed an equally high classification performance of the clinical models of the PPMI sample alone compared to the combined samples models, meaning that the combined sample is not biased by the representation of only the one class in the AmsterdamUMC cohort.

**Table S16 Cross-validated performance evaluation of ICD vs non-ICD classification models in the PPMI dataset with 4th year prediction cut-off**

| Feature subset | Model | Accuracy | Balanced accuracy | Sensitivity | Specificity | AUC |
|---|---|---|---|---|---|---|
| **Clinical features** | RF | 0.66 ± 0.08 | 0.65 ± 0.08 | 0.63 ± 0.15 | 0.67 ± 0.12 | 0.73 ± 0.08 |
| | LR | 0.67 ± 0.09 | 0.67 ± 0.09 | 0.69 ± 0.17 | 0.65 ± 0.17 | 0.72 ± 0.09 |
| | GBC | 0.64 ± 0.08 | 0.62 ± 0.07 | 0.53 ± 0.11 | 0.71 ± 0.12 | 0.67 ± 0.07 |
| | MLP | 0.67 ± 0.09 | 0.65 ± 0.09 | 0.52 ± 0.13 | 0.78 ± 0.13 | 0.72 ± 0.09 |
| **Clinical + Genetics features** | RF | 0.69 ± 0.09 | 0.68 ± 0.09 | 0.63 ± 0.17 | 0.73 ± 0.13 | 0.73 ± 0.09 |
| | LR | 0.67 ± 0.10 | 0.67 ± 0.10 | 0.65 ± 0.19 | 0.69 ± 0.14 | 0.71 ± 0.10 |
| | GBC | 0.68 ± 0.08 | 0.65 ± 0.08 | 0.55 ± 0.15 | 0.76 ± 0.10 | 0.70 ± 0.08 |
| | MLP | 0.68 ± 0.08 | 0.65 ± 0.08 | 0.53 ± 0.15 | 0.77 ± 0.10 | 0.70 ± 0.08 |
| **Clinical + Genetics + Radiomics features** | RF | 0.69 ± 0.08 | 0.66 ± 0.09 | 0.57 ± 0.15 | 0.76 ± 0.09 | 0.74 ± 0.09 |
| | LR | 0.67 ± 0.09 | 0.66 ± 0.10 | 0.62 ± 0.18 | 0.70 ± 0.10 | 0.72 ± 0.10 |
| | GBC | 0.69 ± 0.09 | 0.67 ± 0.10 | 0.56 ± 0.17 | 0.78 ± 0.11 | 0.74 ± 0.10 |
| | MLP | 0.60 ± 0.10 | 0.58 ± 0.09 | 0.49 ± 0.19 | 0.66 ± 0.18 | 0.61 ± 0.09 |

Abbreviations: AUC = Area Under Curve, RF = Random Forest, LR = Logistic Regression, GB = Gradient Boosting, MLP = Multi-Layer Perceptron

**B. Comparison ICD versus No ICD with adding medication use at time of ICD development as predictor**

Since DRT is considered the primary risk factor of ICD development we additionally tested the models with using medication received at the time of ICD positive screening as predictor. Specifically, we included the LEDD received at follow-up, as well as nominal values (yes/no) regarding usage of levodopa, dopamine agonists, MAO-B inhibitors and COM-T inhibitors. Participants with missing values were excluded from the analysis. The models including Clinical + Medication data present a significantly higher performance than the models using only clinical data (AUC up to 0.71±0.06 vs. 0.67±0.06; Table S17). The permutation feature importance method indicated Age of onset (MDA=0.04±0.04), Anxiety severity(MDA=0.03±0.04) and LEDD at follow-up (MDA=0.02±0.04) to being the most important for the predictions.

**Table S17 Cross-validated performance evaluation of ICD vs non-ICD classification models in the full combined dataset (PPMI+AmsterdamUMC)**

| Feature subset | Model | Accuracy | Balanced accuracy | Sensitivity | Specificity | AUC |
|---|---|---|---|---|---|---|
| **Clinical features** | RF | 0.63 ± 0.05 | 0.61 ± 0.05 | 0.56 ± 0.10 | 0.67 ± 0.08 | 0.66 ± 0.05 |
| | LR | 0.63 ± 0.06 | 0.62 ± 0.06 | 0.58 ± 0.09 | 0.67 ± 0.08 | 0.67 ± 0.06 |
| | GBC | 0.59 ± 0.06 | 0.56 ± 0.06 | 0.43 ± 0.10 | 0.69 ± 0.08 | 0.60 ± 0.07 |
| | MLP | 0.63 ± 0.04 | 0.53 ± 0.04 | 0.28 ± 0.08 | 0.79 ± 0.06 | 0.66 ± 0.07 |
| **Clinical + Medication features** | RF | 0.65 ± 0.07 | 0.64 ± 0.07 | 0.57 ± 0.10 | 0.71 ± 0.09 | 0.69 ± 0.07 |
| | LR | 0.65 ± 0.06 | 0.64 ± 0.06 | 0.62 ± 0.10 | 0.67 ± 0.09 | 0.71 ± 0.06 |
| | GBC | 0.59 ± 0.06 | 0.56 ± 0.06 | 0.42 ± 0.12 | 0.69 ± 0.08 | 0.59 ± 0.07 |
| | MLP | 0.65 ± 0.05 | 0.55 ± 0.06 | 0.18 ± 0.18 | 0.93 ± 0.09 | 0.70 ± 0.08 |
| **Clinical + Medication + Radiomics features** | RF | 0.62 ± 0.06 | 0.60 ± 0.06 | 0.52 ± 0.10 | 0.69 ± 0.09 | 0.65 ± 0.06 |
| | LR | 0.61 ± 0.07 | 0.60 ± 0.07 | 0.59 ± 0.11 | 0.62 ± 0.09 | 0.65 ± 0.07 |
| | GBC | 0.61 ± 0.06 | 0.58 ± 0.06 | 0.45 ± 0.12 | 0.70 ± 0.09 | 0.58 ± 0.06 |
| | MLP | 0.59 ± 0.08 | 0.53 ± 0.06 | 0.31 ± 0.26 | 0.76 ± 0.25 | 0.59 ± 0.06 |

Abbreviations: AUC = Area Under Curve, RF = Random Forest, LR = Logistic Regression, GB = Gradient Boosting, MLP = Multi-Layer Perceptron

#### C. Comparison of PD versus Healthy controls

We aimed to check the quality of the extracted radiomic and deep features. To this end, we evaluated ML models to diagnose PD vs HC, which is the main clinical utility of DAT-SPECT in PD, and previously reported ML models have shown excellent performance (16). We used the processed imaging data of the pre-registered analysis (N=378) and also included the HC imaging data of the PPMI (N=180). We applied the same pre-processing and feature extraction steps on the HC sample. We trained three RF models DAT-SPECT using as predictors: only SBRs, only radiomic features and only latent features, respectively. The models were tested with 10x5 fold cross-validation and showed excellent accuracy. Results are shown in Table S18.

**Table S18 Cross-validated performance evaluation of PD vs healthy controls.**

| Feature subset | Model | Accuracy | Sensitivity | Specificity | AUC |
|---|---|---|---|---|---|
| **SBRs** | RF | 0.95 ± 0.02 | 0.95 ± 0.02 | 0.95 ± 0.02 | 0.98 ± 0.02 |
| **Radiomic features** | RF | 0.94 ± 0.02 | 0.93 ± 0.02 | 0.97 ± 0.02 | 0.99 ± 0.02 |
| **Latent features*** | RF | 0.95 ± 0.02 | 0.94 ± 0.02 | 0.98 ± 0.02 | 0.99 ± 0.02 |

*Latent features were extracted with the 3D autoencoder network.

**REFERENCES**


1. Vriend C, Nordbeck AH, Booij J, van der Werf YD, Pattij T, Voorn P, et al. Reduced dopamine transporter binding predates impulse control disorders in Parkinson's disease. Mov Disord. 2014;29(7):904-11.
2. Trujillo JP, Gerrits NJ, Vriend C, Berendse HW, van den Heuvel OA, van der Werf YD. Impaired planning in Parkinson's disease is reflected by reduced brain activation and connectivity. Hum Brain Mapp. 2015;36(9):3703-15.
3. Laansma MA, Bright JK, Al-Bachari S, Anderson TJ, Ard T, Assogna F, et al. International Multicenter Analysis of Brain Structure Across Clinical Stages of Parkinson's Disease. Mov Disord. 2021;36(11):2583-94.
4. Hentz JG, Mehta SH, Shill HA, Driver-Dunckley E, Beach TG, Adler CH. Simplified conversion method for unified Parkinson's disease rating scale motor examinations. Mov Disord. 2015;30(14):1967-70.
5. Beck AT, Ward CH, Mendelson M, Mock J, Erbaugh J. An inventory for measuring depression. Archives of general psychiatry. 1961;4:561-71.
6. Yesavage JA, Sheikh JI. 9/Geriatric depression scale (GDS) recent evidence and development of a shorter version. Clinical gerontologist. 1986;5(1-2):165-73.
7. Maust D, Cristancho M, Gray L, Rushing S, Tjoa C, Thase ME. Psychiatric rating scales. Handb Clin Neurol. 2012;106:227-37.
8. Spielberger CD, Goruch R, Lushene R, Vagg P, Jacobs G. Manual for the state-trait inventory STAI (form Y). Mind Garden, Palo Alto, CA, USA. 1983.
9. van Steenoven I, Aarsland D, Hurtig H, Chen-Plotkin A, Duda JE, Rick J, et al. Conversion between mini-mental state examination, montreal cognitive assessment, and dementia rating scale-2 scores in Parkinson's disease. Mov Disord. 2014;29(14):1809-15.
10. Rahmim A, Salimpour Y, Jain S, Blinder SA, Klyuzhin IS, Smith GS, et al. Application of texture analysis to DAT SPECT imaging: Relationship to clinical assessments. Neuroimage Clin. 2016;12:e1-e9.
11. Rahmim A, Huang P, Shenkov N, Fotouhi S, Davoodi-Bojd E, Lu L, et al. Improved prediction of outcome in Parkinson's disease using radiomics analysis of longitudinal DAT SPECT images. Neuroimage Clin. 2017;16:539-44.
12. van Griethuysen JJM, Fedorov A, Parmar C, Hosny A, Aucoin N, Narayan V, et al. Computational Radiomics System to Decode the Radiographic Phenotype. Cancer Res. 2017;77(21):e104-e7.
13. Martini A, Dal Lago D, Edelstyn NMJ, Salgarello M, Lugoboni F, Tamburin S. Dopaminergic Neurotransmission in Patients With Parkinson's Disease and Impulse Control Disorders: A Systematic Review and Meta-Analysis of PET and SPECT Studies. Frontiers in neurology. 2018;9:1018.
14. Smith KM, Xie SX, Weintraub D. Incident impulse control disorder symptoms and dopamine transporter imaging in Parkinson disease. J Neurol Neurosurg Psychiatry. 2016;87(8):864-70.
15. Hosseinzadeh M, Gorji A, Fathi Jouzdani A, Rezaeijo SM, Rahmim A, Salmanpour MR. Prediction of Cognitive Decline in Parkinson's Disease Using Clinical and DAT SPECT Imaging Features, and Hybrid Machine Learning Systems. Diagnostics (Basel). 2023;13(10).
16. Shiiba T, Takano K, Takaki A, Suwazono S. Dopamine transporter single-photon emission computed tomography-derived radiomics signature for detecting Parkinson's disease. EJNMMI Res. 2022;12(1):39.
